## Supplementary Figures for "Genome-wide association analysis of plasma lipidome identifies 495 genetic associations"

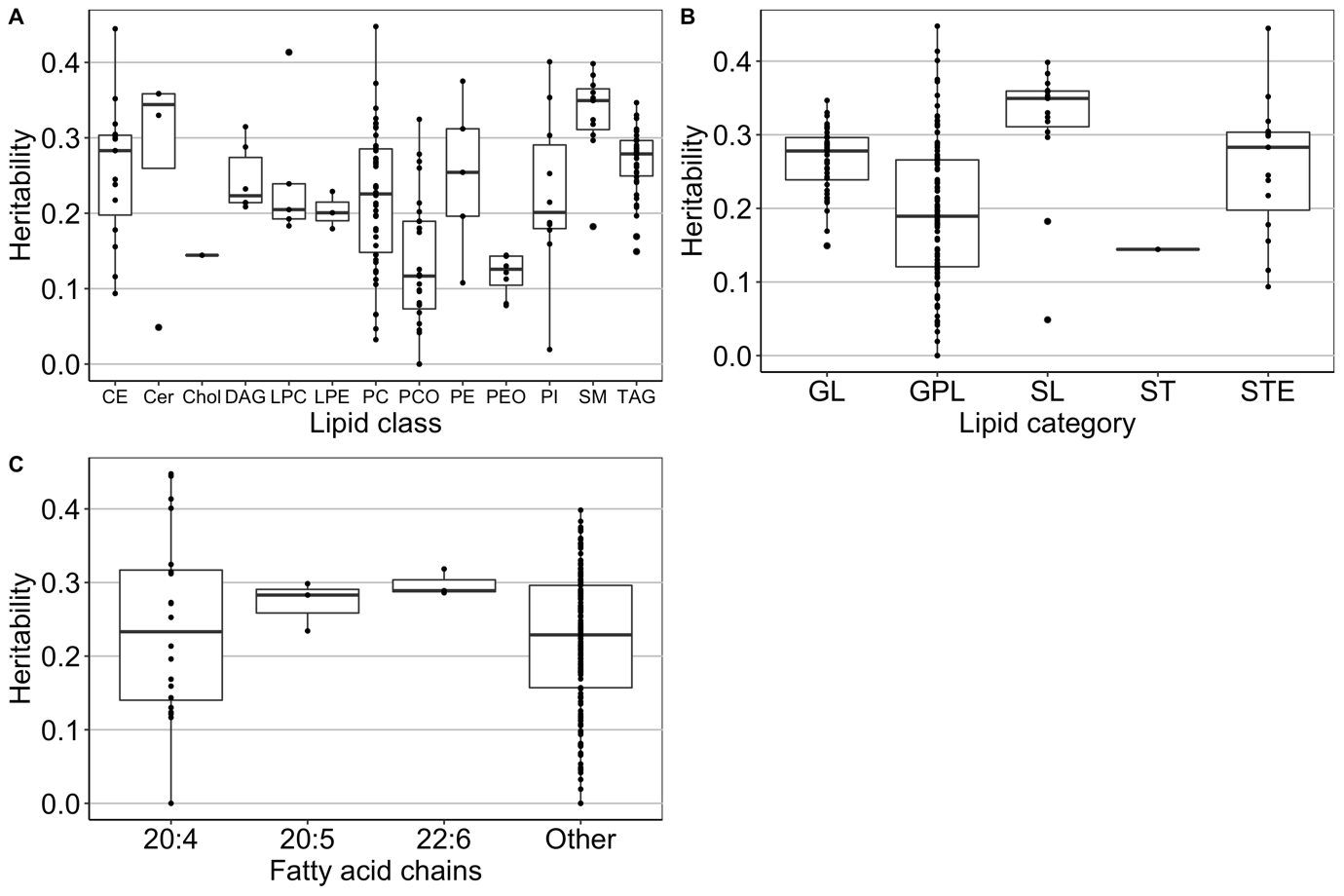

**Supplementary Figure 1.** Box plots of heritability estimates between species of different lipid classes (A), lipid categories (B) and species containing specific fatty acid chains (C). The boxplots depict the median (middle line), interquartile range (IQR) defining the bounds of the box, and whiskers extending to the largest/smallest values no further than 1.5 times the IQR. Data beyond the end of whiskers are plotted individually.

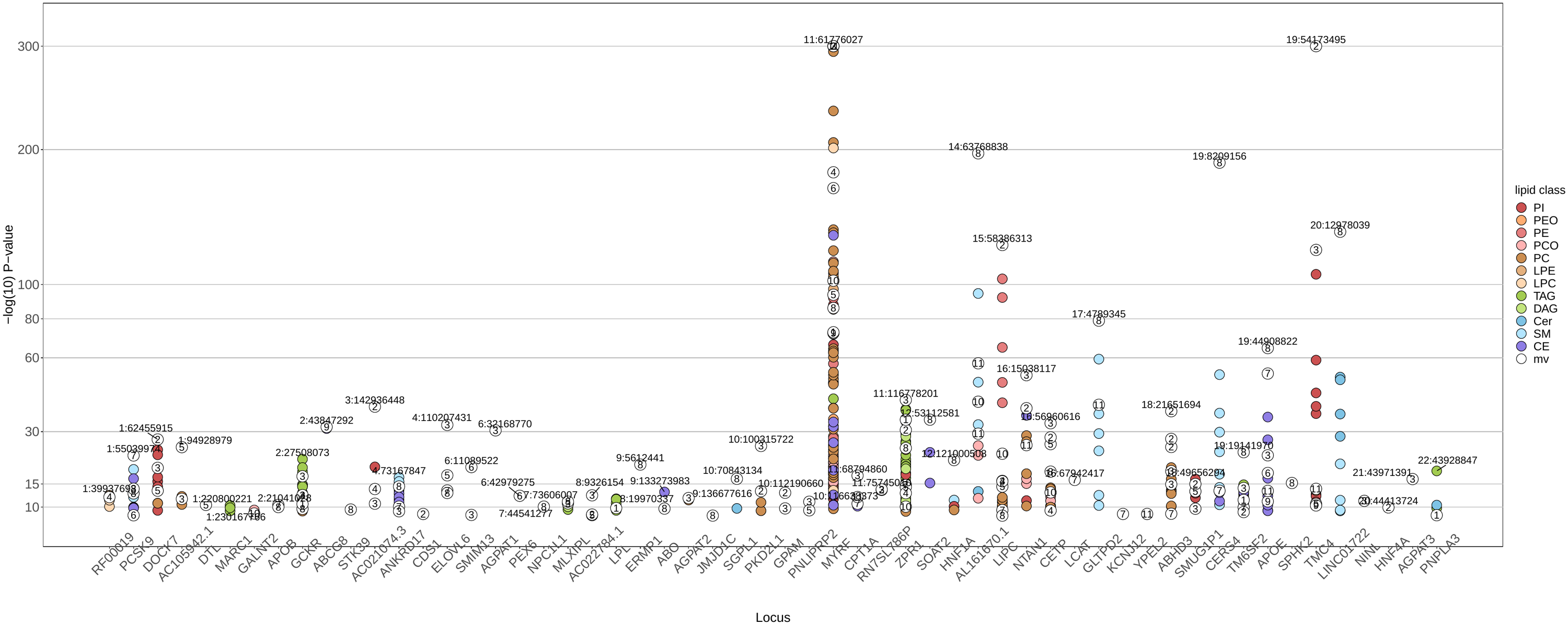

**Supplementary Figure 2. Univariate and multivariate P-values for loci reaching Bonferroni-corrected threshold.** Loci are colored by lipid class in univariate analysis and labeled by cluster number from multivariate analysis. The top variant for each locus is annotated by chromosome:base pair position (GRCh38). Lipid class names are listed in Figure 1. Y-axis is capped at 300.

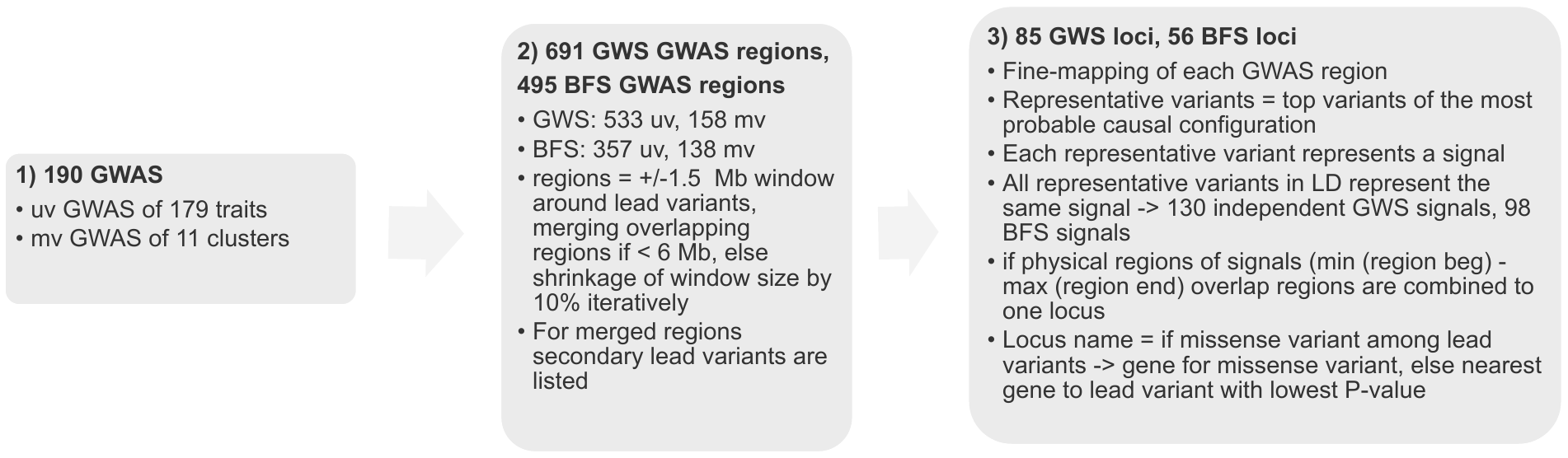

**Supplementary Figure 3. Flowchart of the locus definition process.** Numbers of GWAS regions and loci reaching the genome-wide significance (GWS) threshold and the Bonferroni-corrected significance (BFS) threshold are given.

**Supplementary Figure 4. Locus definition process for locus *LPL*.** 1. P-values of variants in the 3 Mb GWAS region around lead variant (colored red) for TAG 52:4;0. Not shown here: overlapping GWAS regions are merged (see Methods). 2. P-values of variants in 0.2 Mb window around the lead variant for TAG 52:4;0. Representative variants are colored (red and blue), the red representative variant is also the lead variant. 3. P-values of variants in 0.2 Mb window around the lead variant for TAG 52:4;0 and P-values in the same region for TAG 56:5;0. The representative variant for TAG 56:5;0 (blue) is in LD with the blue representative variant for TAG 52:4;0 and therefore represents the same signal (signal 2). The position of the representative variant of signal 2 (blue) is marked for both traits. 4. Regions of the signals and the locus.

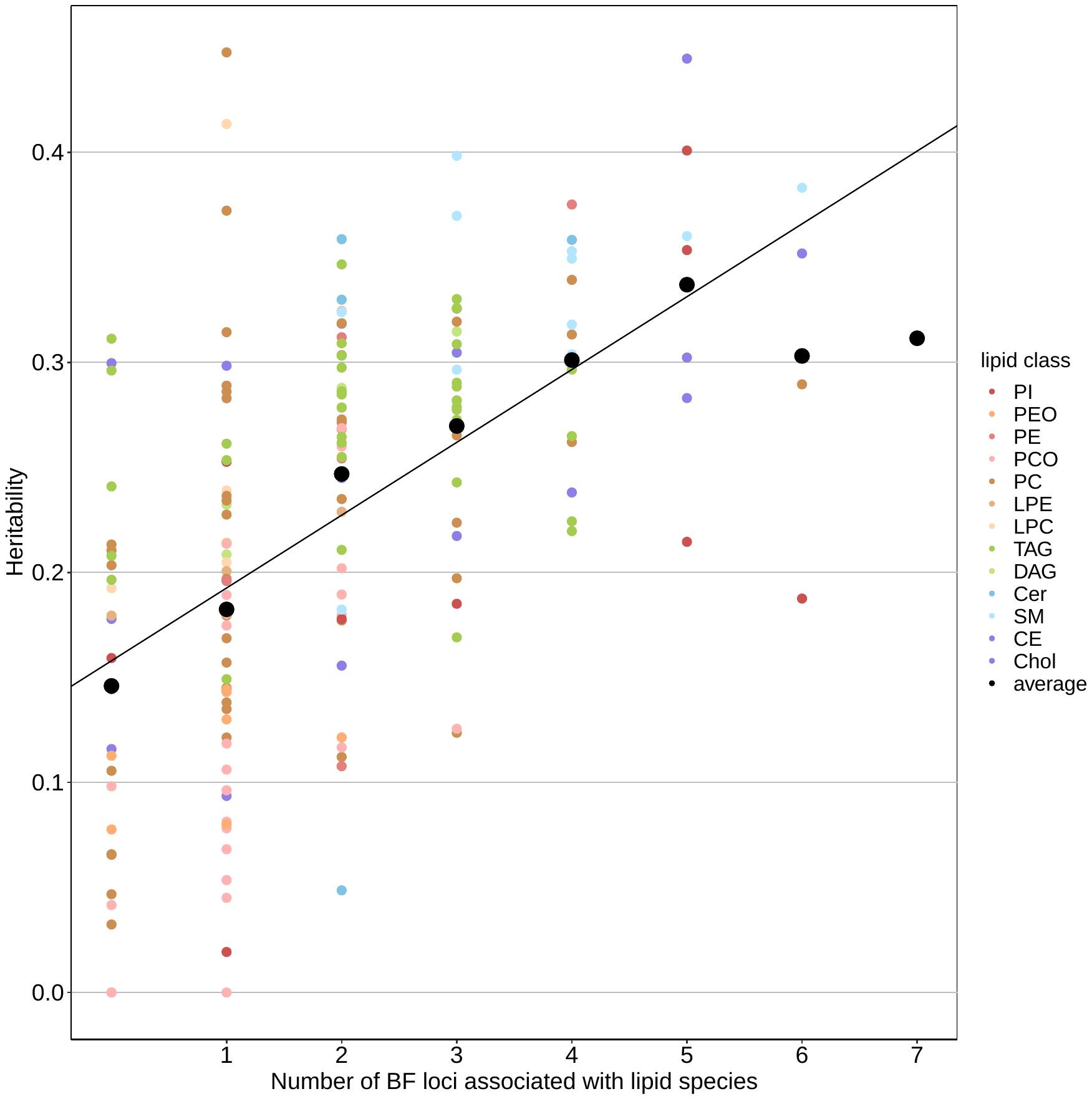

**Supplementary Figure 5. Relationship between heritability estimates and the number of loci reaching the Bonferroni-corrected significance threshold of 7.35e-10 per lipid species.** The black line shows the linear regression fit (intercept=0.157927;se=0.009911, slope=0.034667;se=0.003888) with weights of the linear regression set to 1/se^2^. Larger black dots depict the average estimated heritability and smaller dots depict the individual heritability estimates colored by lipid class.

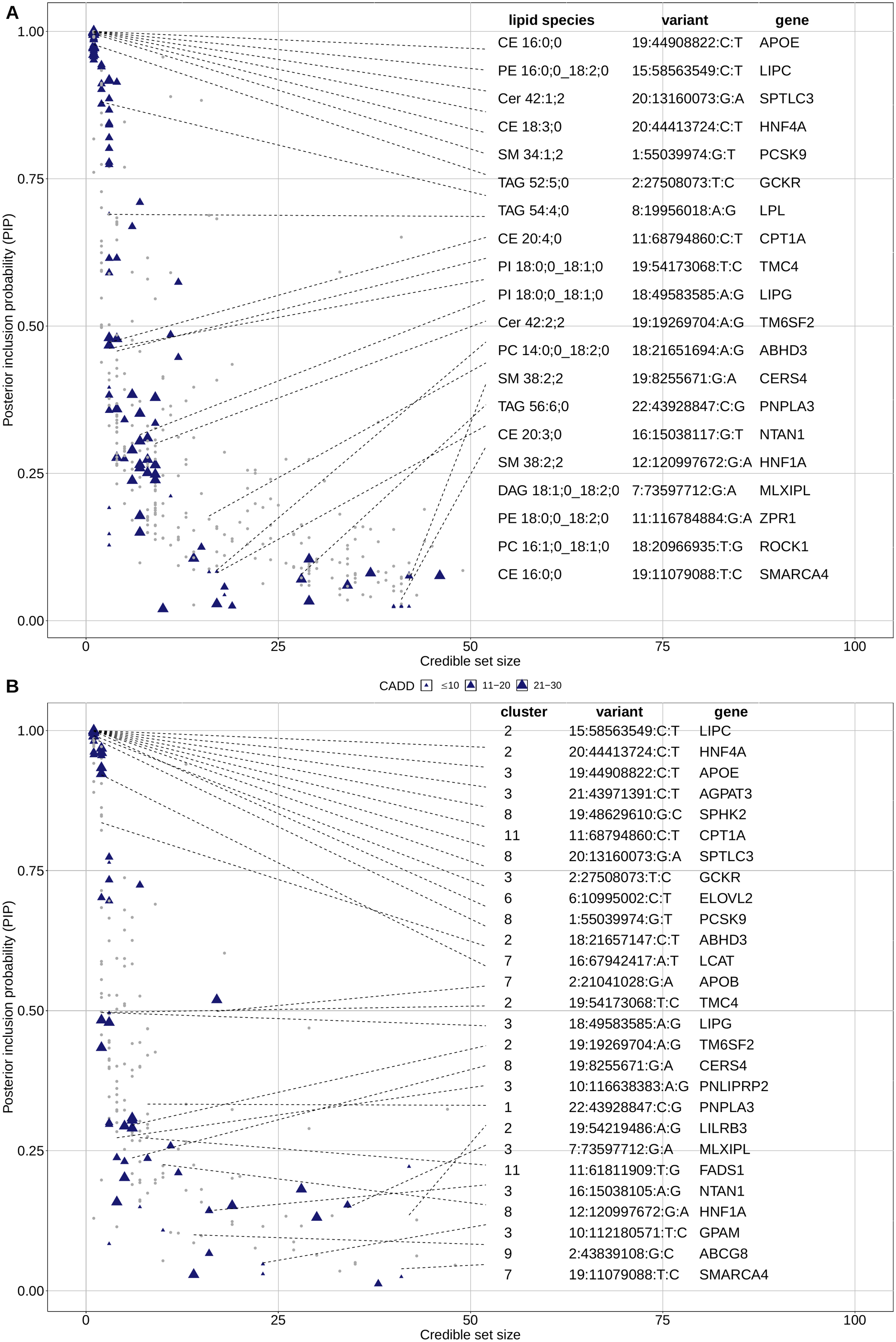

**Supplementary Figure 6. Representative variants of informative 95% credible sets with credible set size < 50 and top PIP > 1%** **of BFS loci in (A) univariate GWAS (B) multivariate GWAS plotted by credible set size against PIP.** Blue triangles are functional variants, other SNPs are shown as gray dots. The size of the triangles is proportional to the variants’ CADD score. For loci for which functional variants were among the representative variants, the functional representative variant with the lowest GWAS P-value is annotated with the top associated trait, variant id, and gene from Variant effect predictor.

A

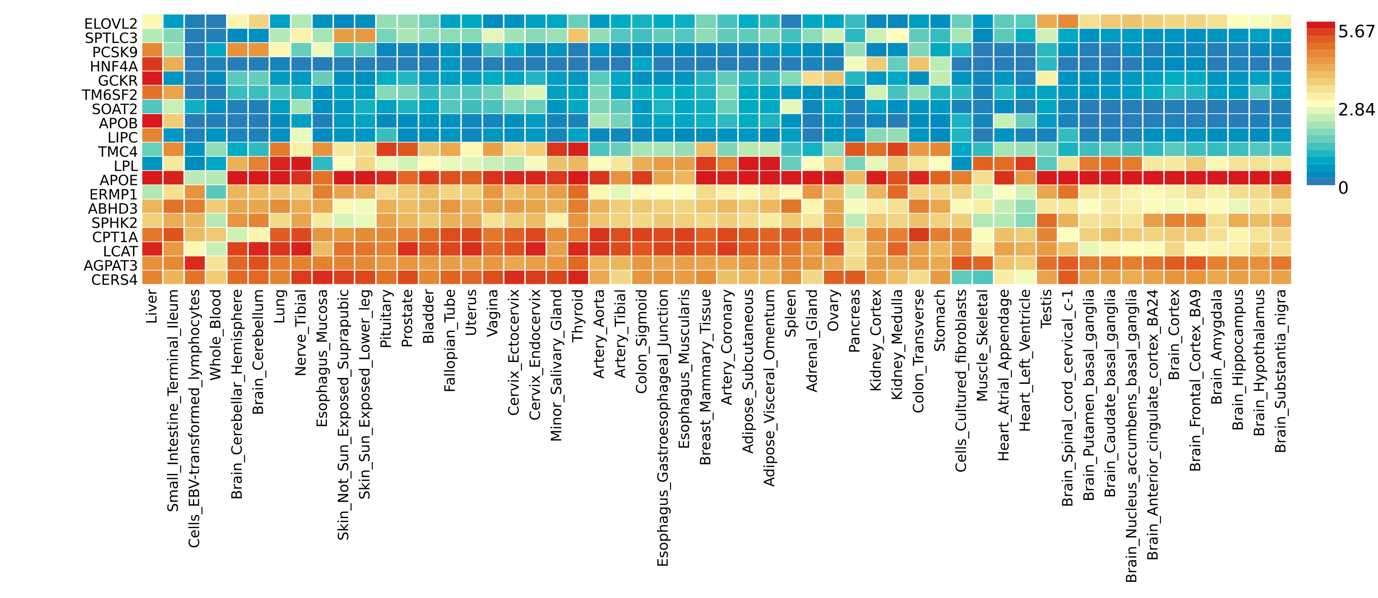

B

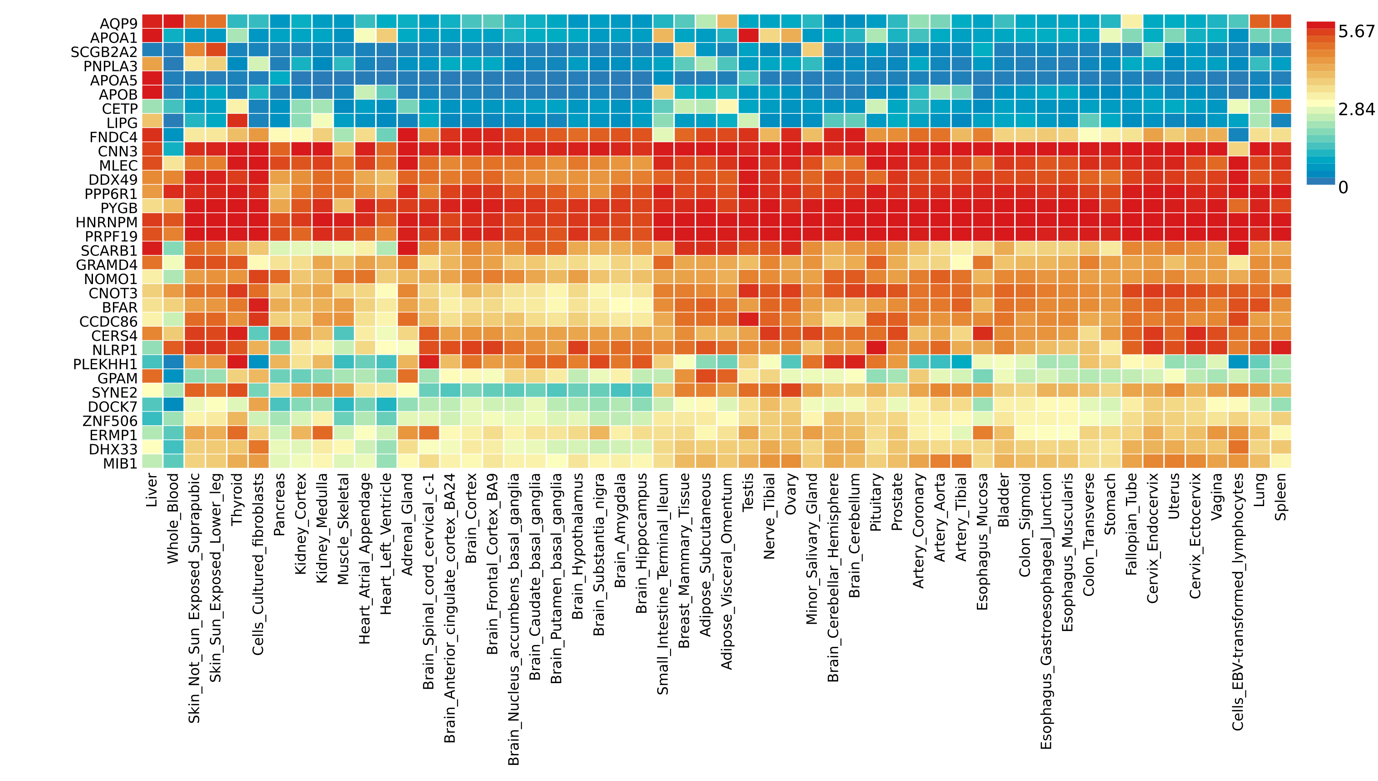

**Supplementary Figure 7. Heatmap of expression level in GTEx tissues of genes prioritized by (A) functional variant approach and (B) FOCUS.** The average log2 transformed expression value is shown.

A

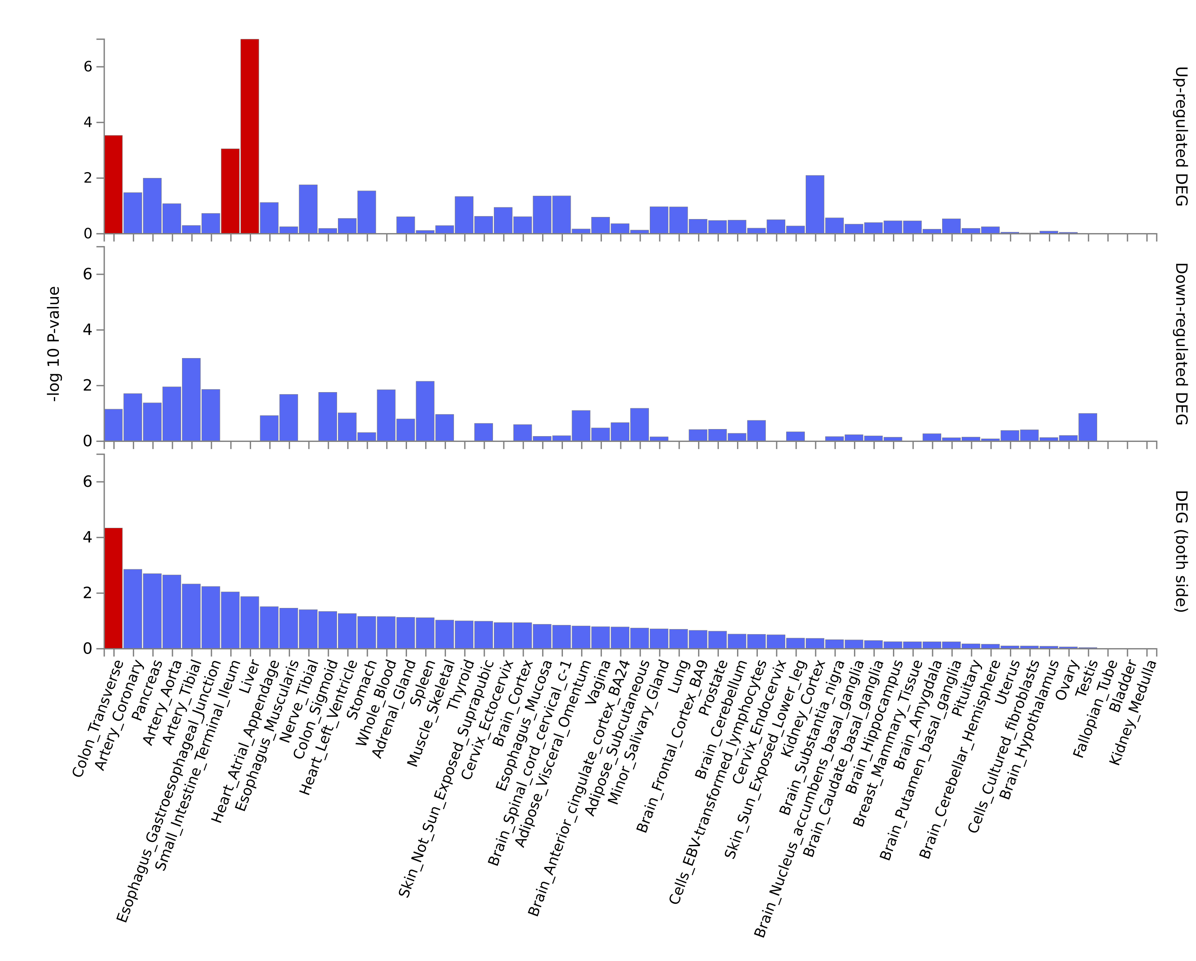

B

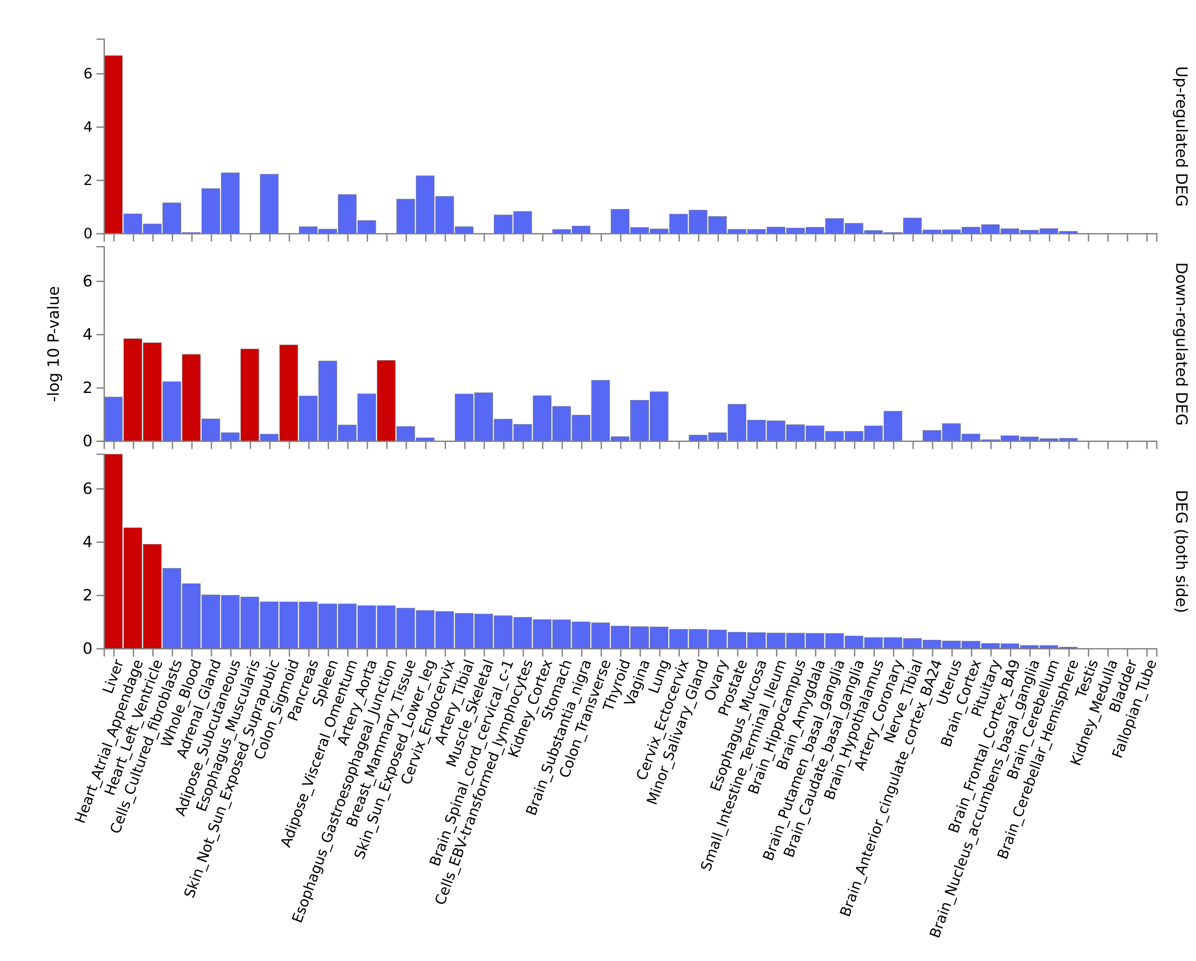

**Supplementary Figure 8. Enrichment of differentially expressed genes (DEG) sets for genes prioritized by (A) functional variant approach and (B) FOCUS in GTEx tissues.** Significantly enriched DEG sets (P$\leq$0.05 corrected for multiple testing) are highlighted in red.

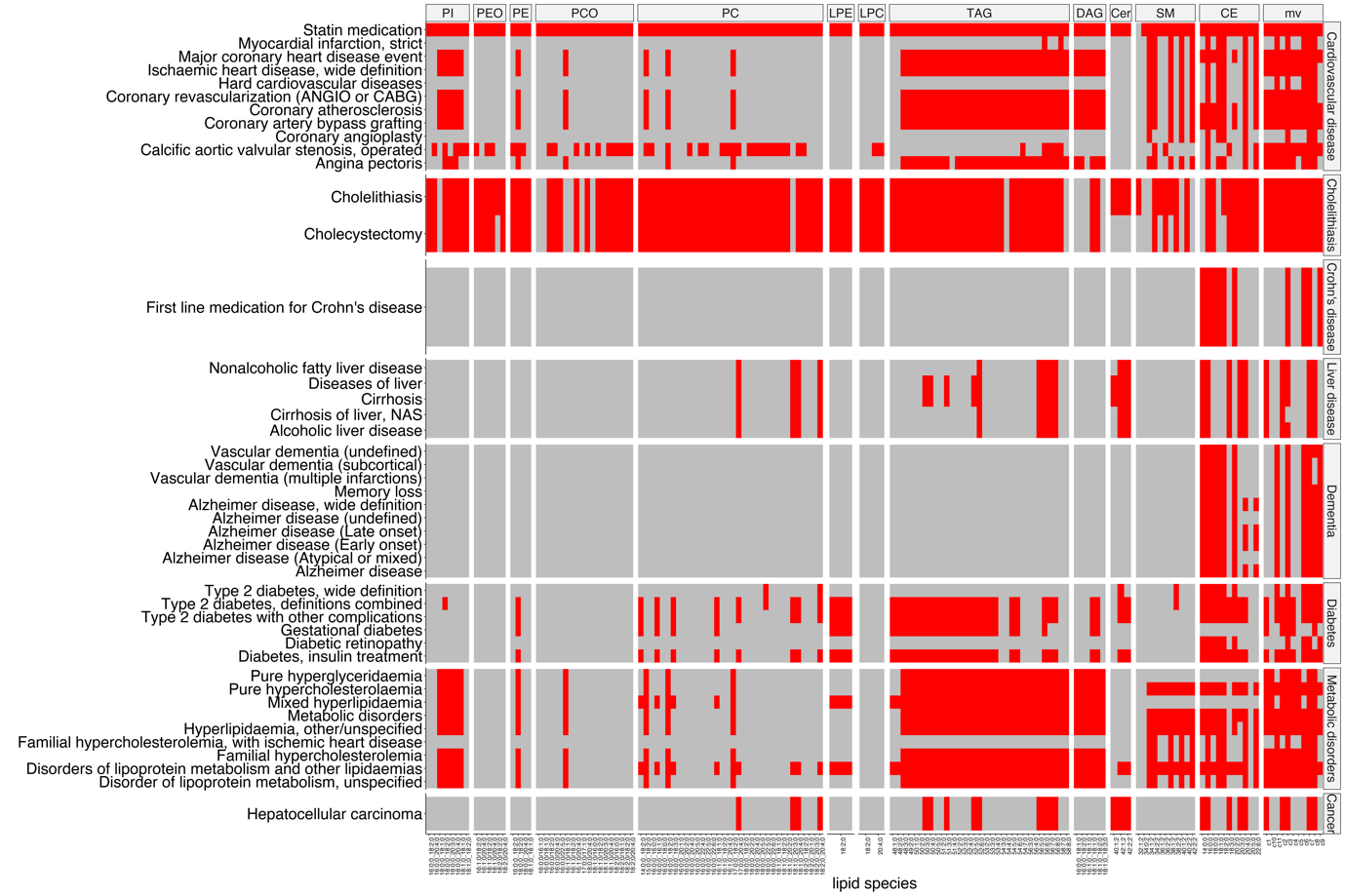

**Supplementary Figure 9. Heatmap of PheWAS associations*.*** Each entry in the heatmap represents a PheWAS endpoint of a disease group (row), which is associated with variants among the lead variants or representative variants of a GWAS (column). Columns are split by lipid classes and rows are split by disease groups. Lead variants reaching the Bonferroni-corrected significance (BFS, uv=7.35e-10, mv=4.55e-9) and representative variants of BFS associations are considered. PheWAS associations reaching BFS (P=5.25e-11) are included. BFS associations are colored red.

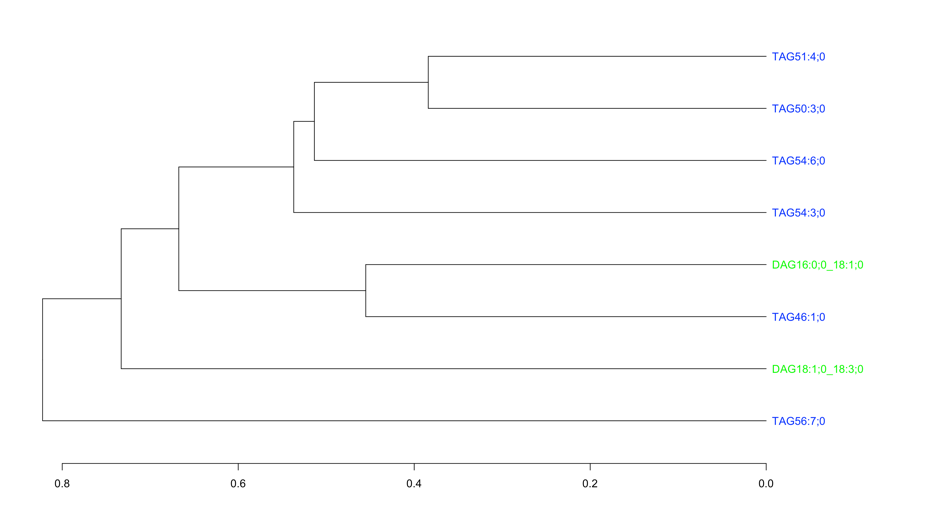

**Supplementary Figure 10. Dendrogram for hierarchical clustering of lipid species of the lipid classes TAG and DAG.** Only lipid species with pairwise correlations < 0.8 were included. All lipid species are contained in cluster1.

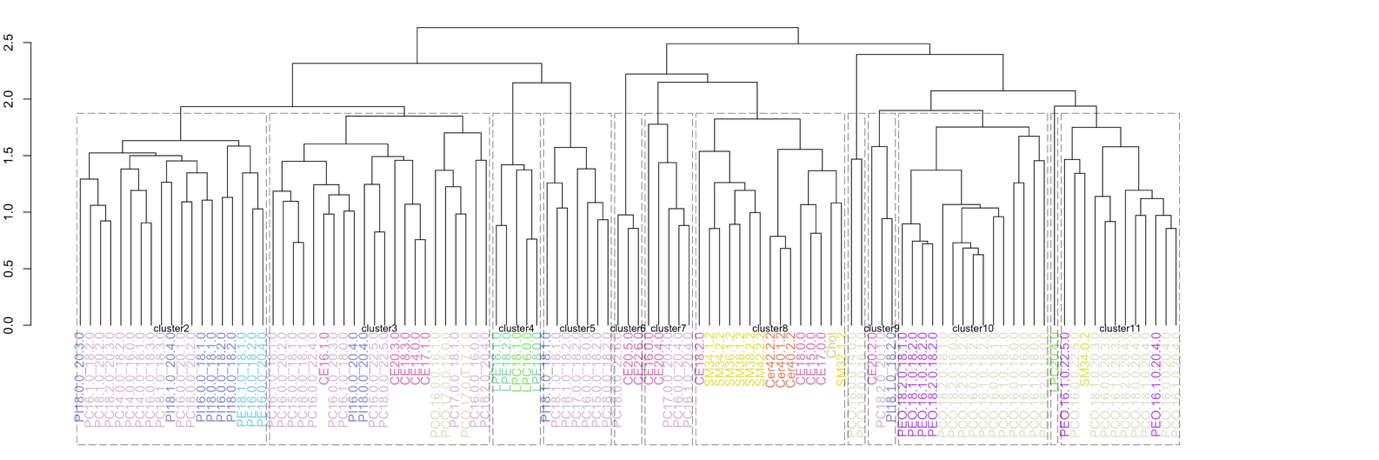

**Supplementary Figure 11. Dendrogram for hierarchical clustering of lipid species of the lipid classes CE, Cer, Chol, LPC, LPE, PC, PCO, PE, PEO, PI and SM.** Only lipid species with pairwise correlations < 0.8 were included. Lipid species contained in a cluster for multivariate analysis are surrounded by rectangles and labelled by cluster name.

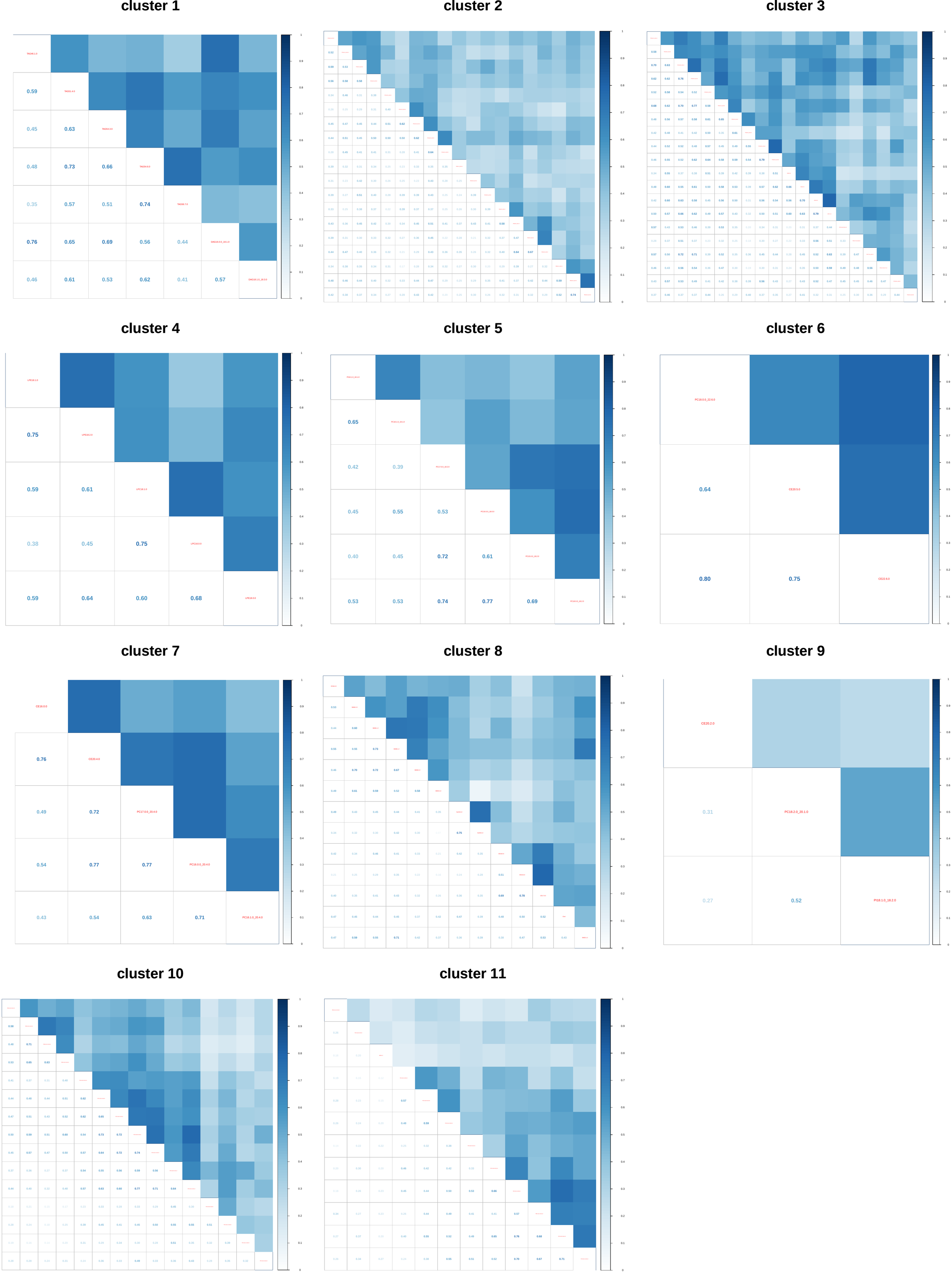

**Supplementary Figure 12. Heatmaps of absolute pairwise Pearson correlations for lipid species included in a specific cluster.**

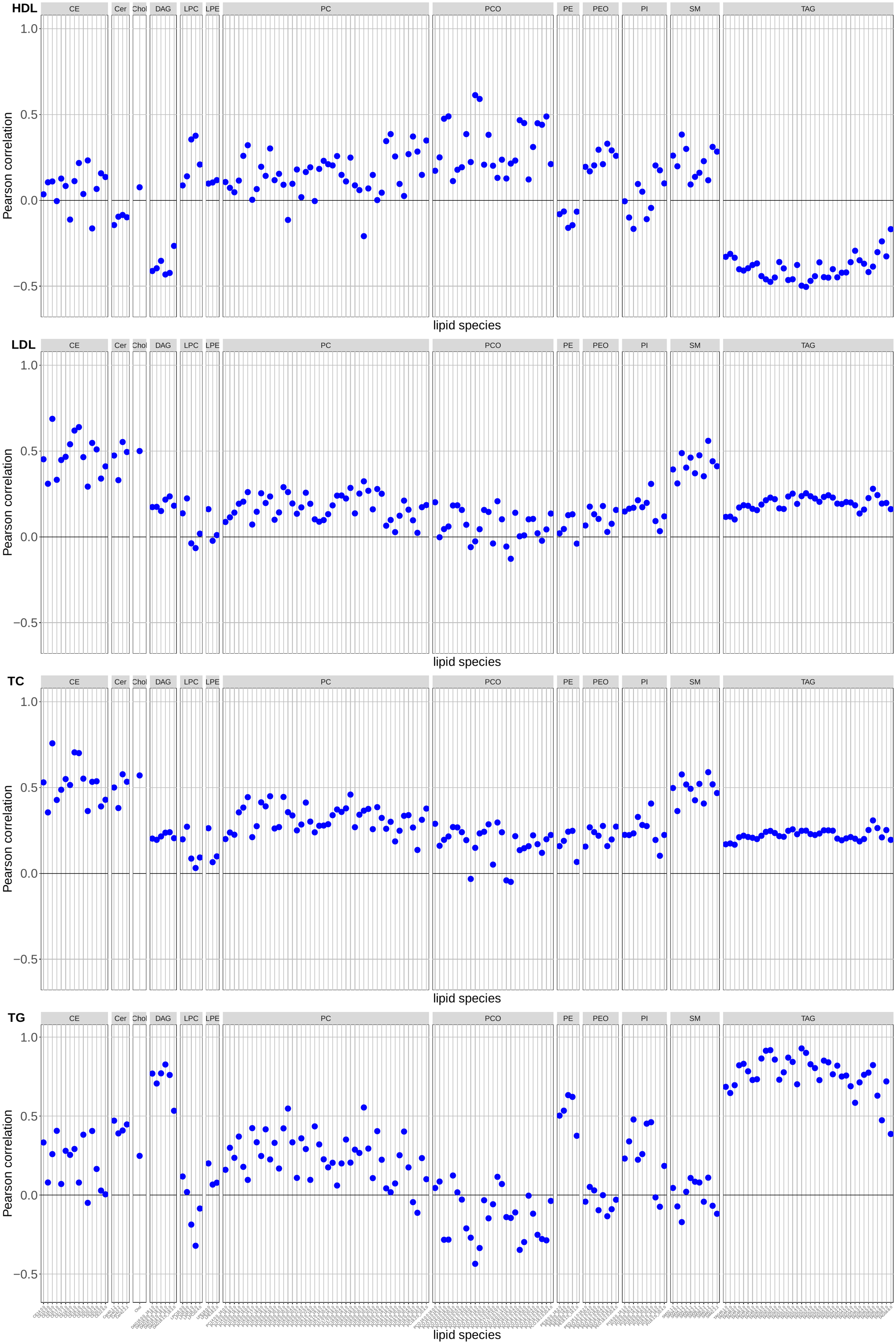

**Supplementary Figure 13. Pairwise Pearson correlations of lipid species and standard lipids.**

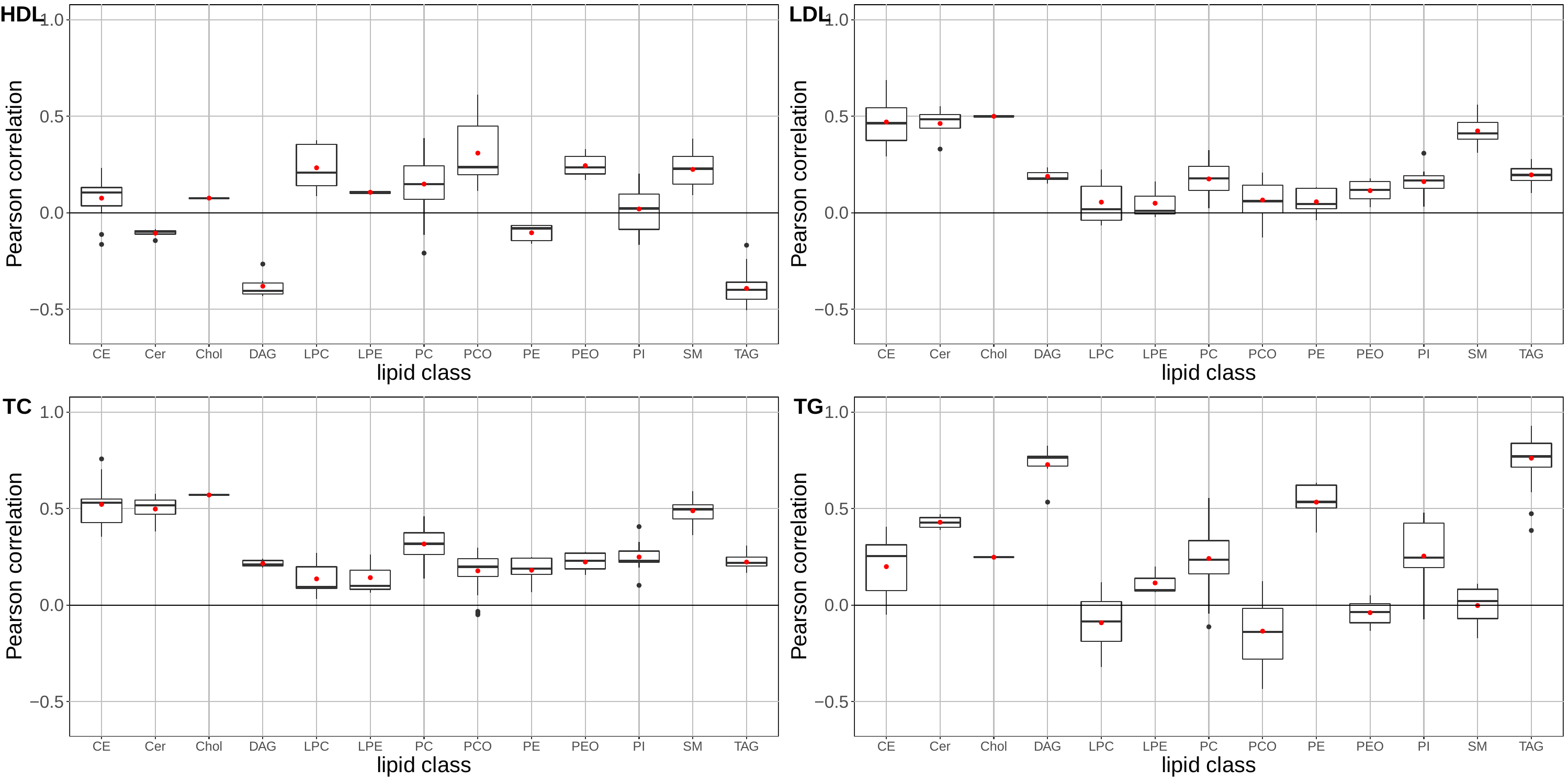

**Supplementary Figure 14. Boxplots of pairwise Pearson correlations of lipid species of different lipid classes and standard lipids.**
