## Supplementary Data 1 for "Genome-wide association analysis of plasma lipidome identifies 495 genetic associations"

**Manhattan plots for lipid classes**

Top associations reaching Bonferroni-corrected P-value threshold of 7.35e-10 are depicted as diamonds and annotated by variant, locus name and lipid species. Known loci are colored yellow and novel loci are colored red. The vertical line indicates the Bonferroni-corrected P-value threshold. Only variants with P-value < 1e-5 are shown. The y-axis is capped at 100.

**CE**

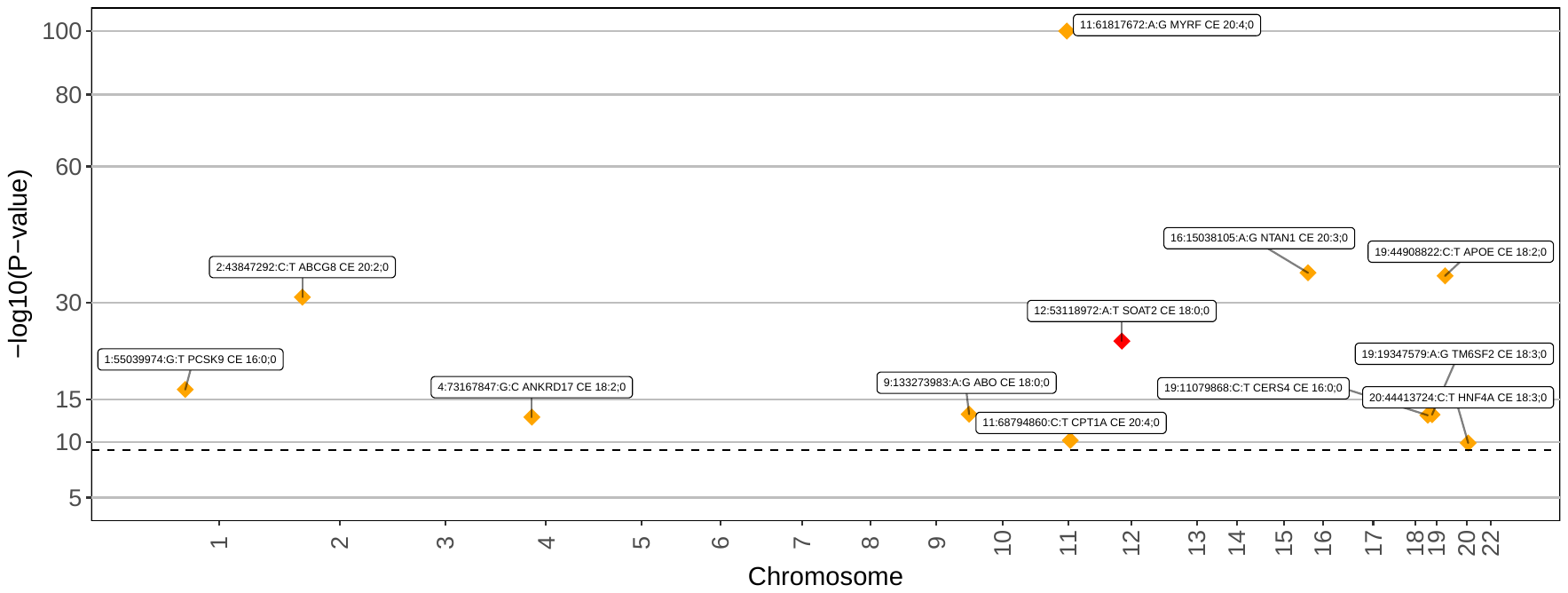

**Cer**

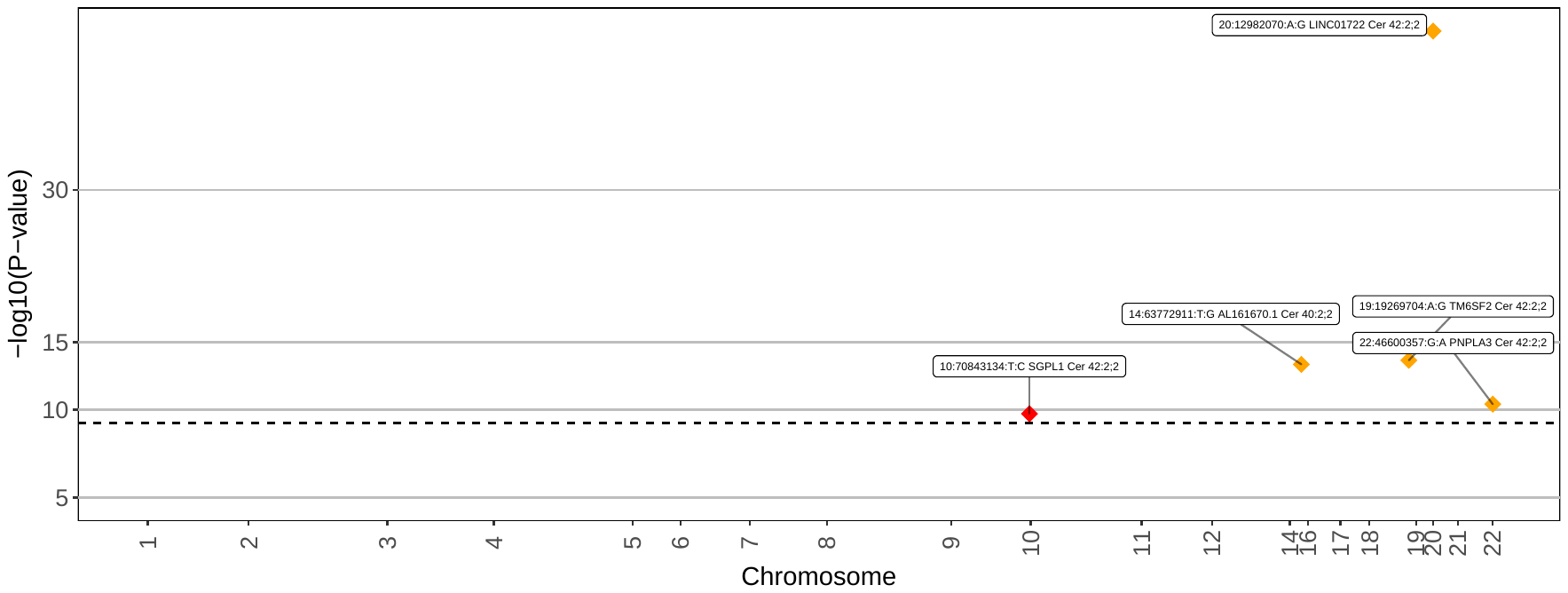

**Chol**

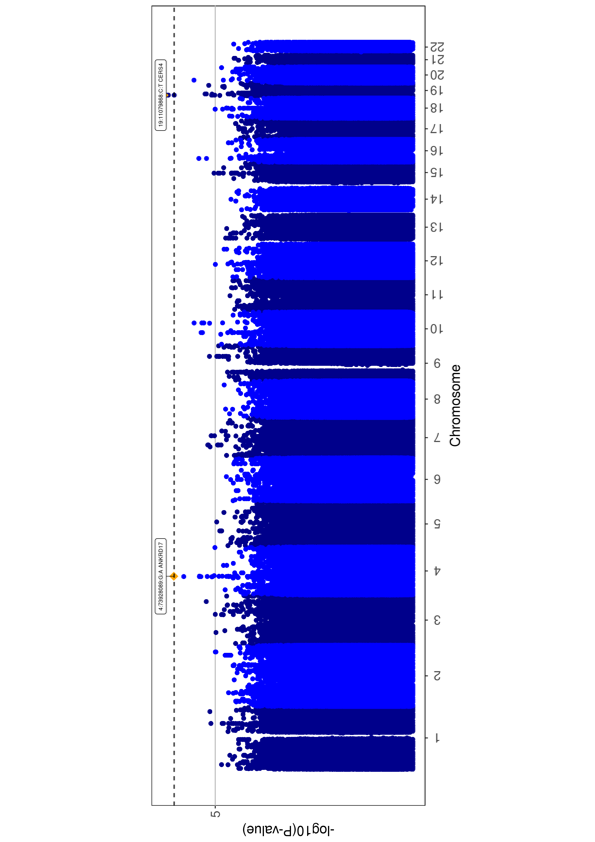

**DAG**

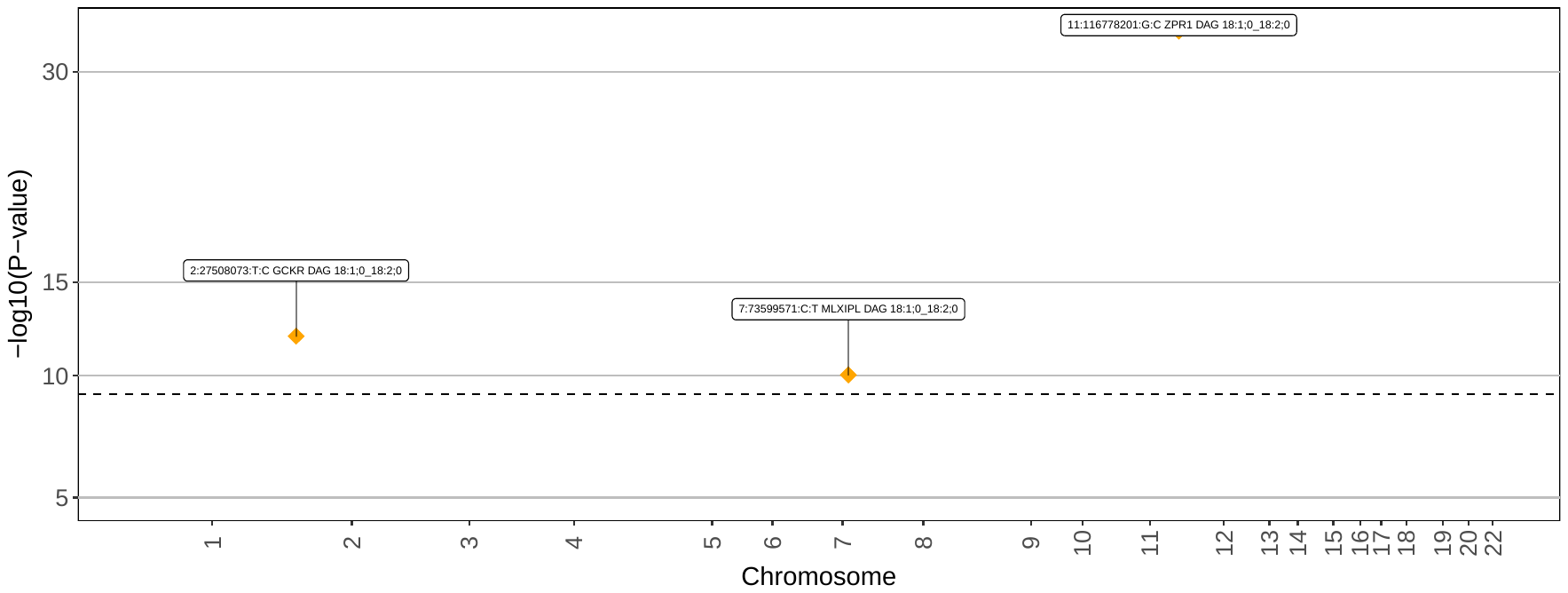

**LPC**

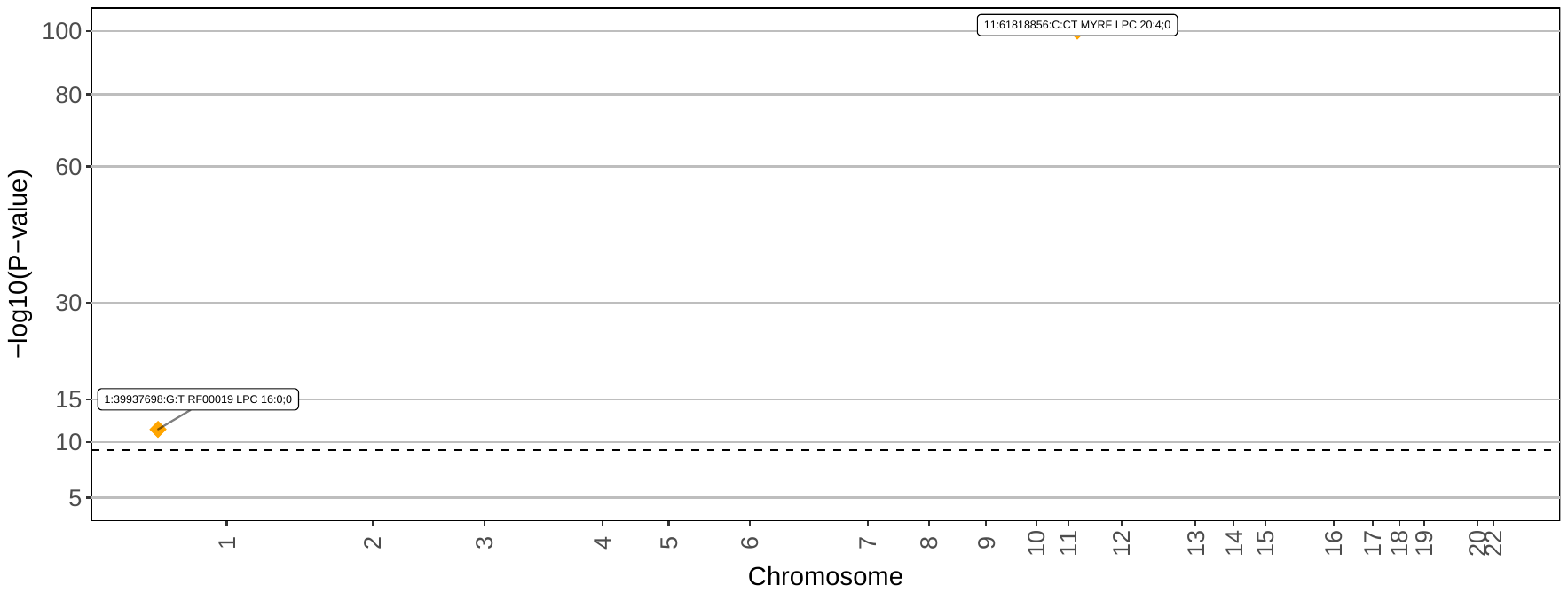

**LPE**

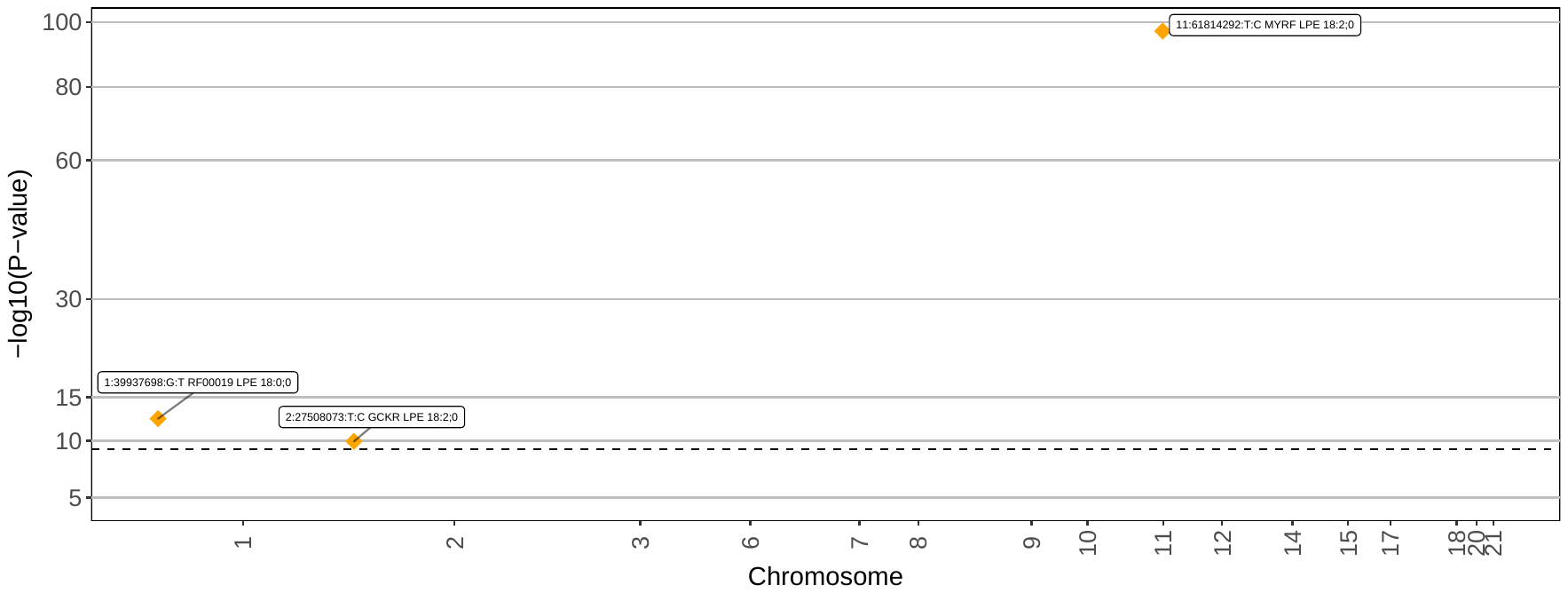

**PC**

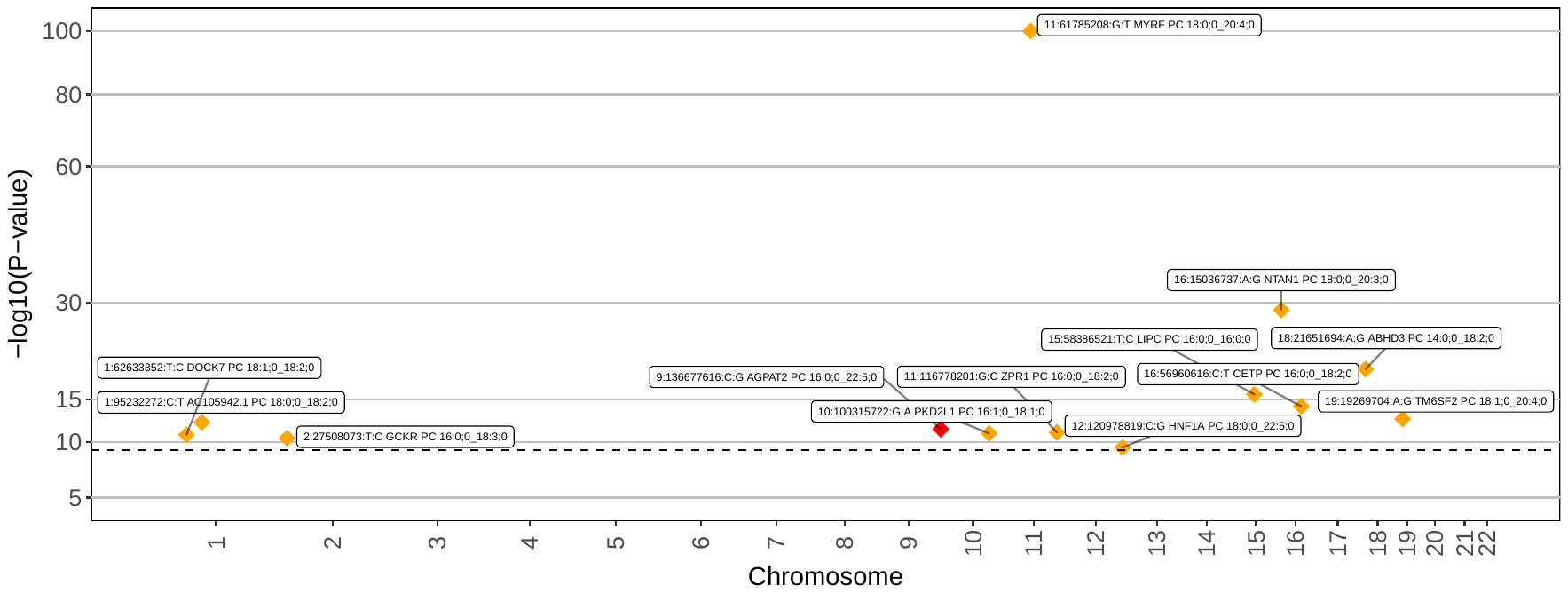

**PCO**

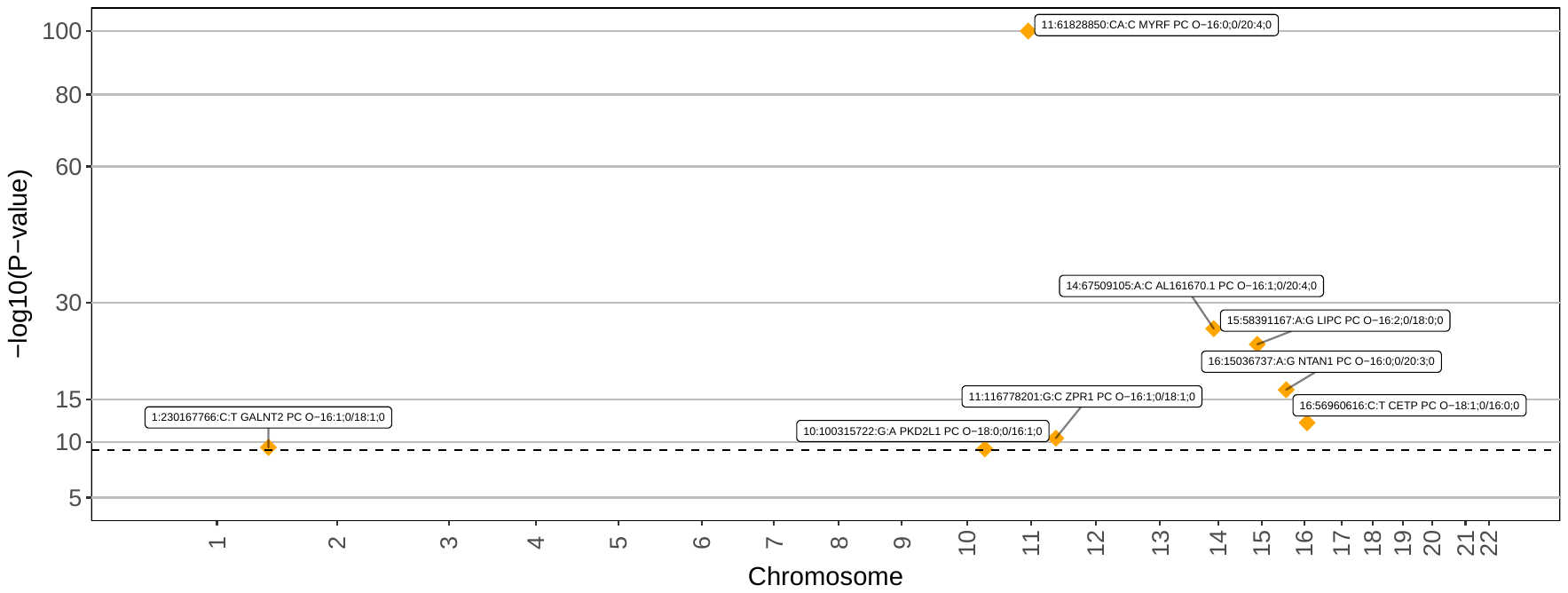

**PE**

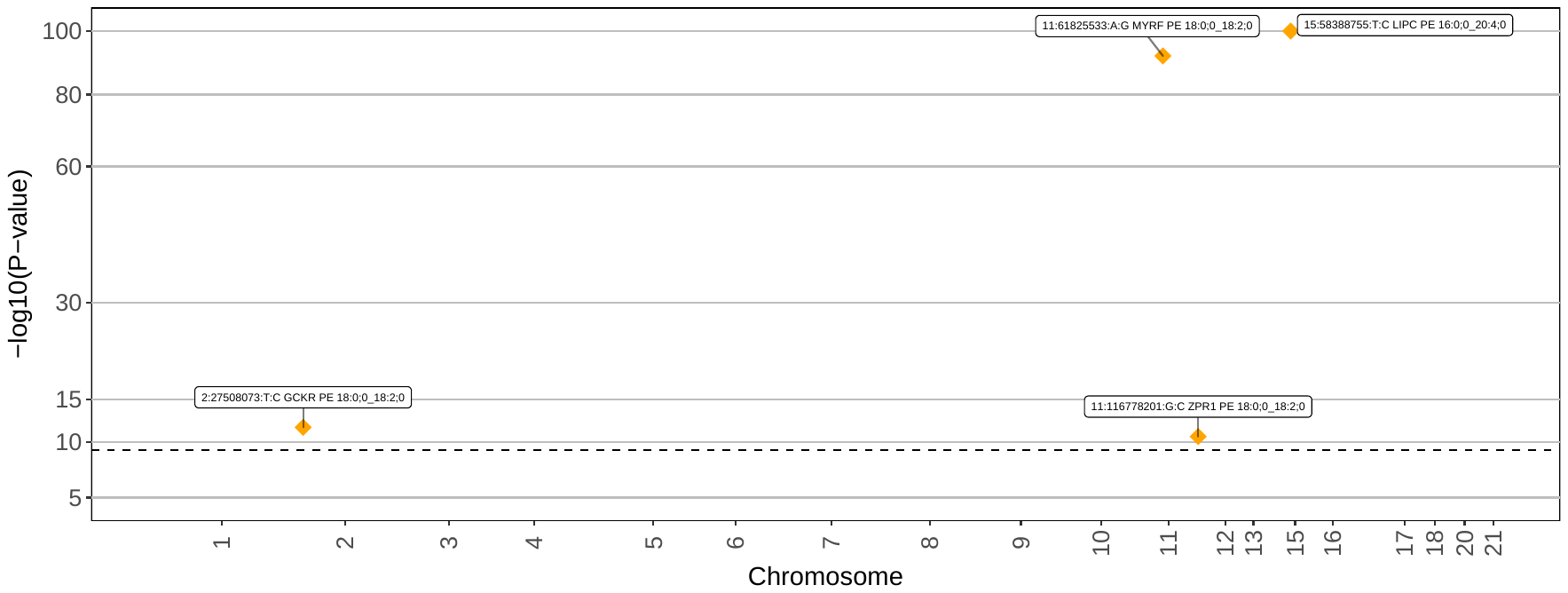

**PEO**

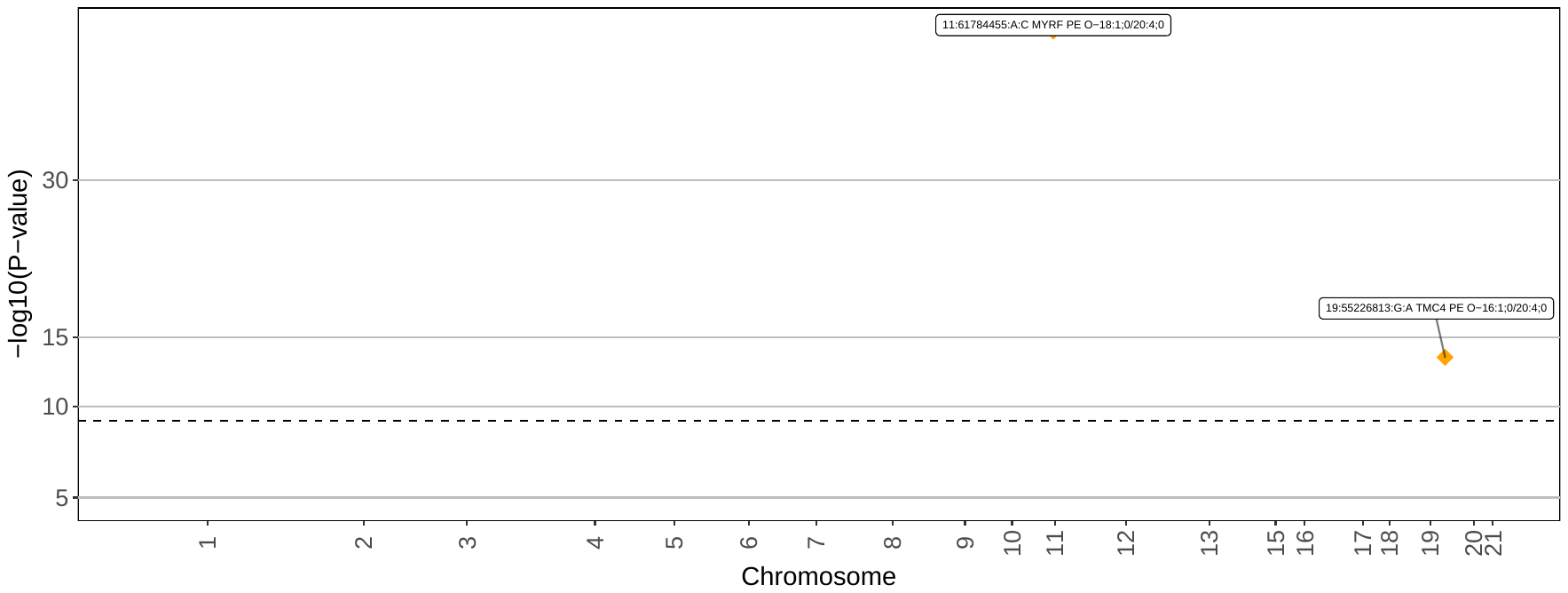

**PI**

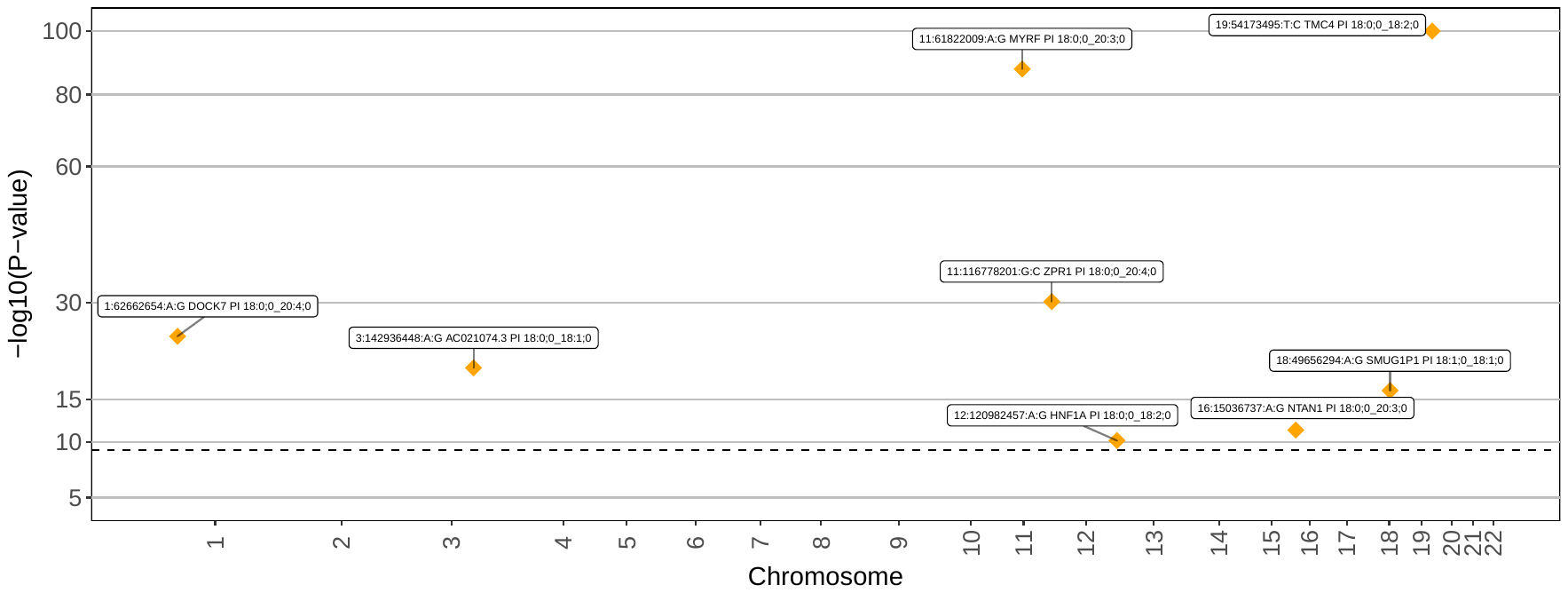

**SM**

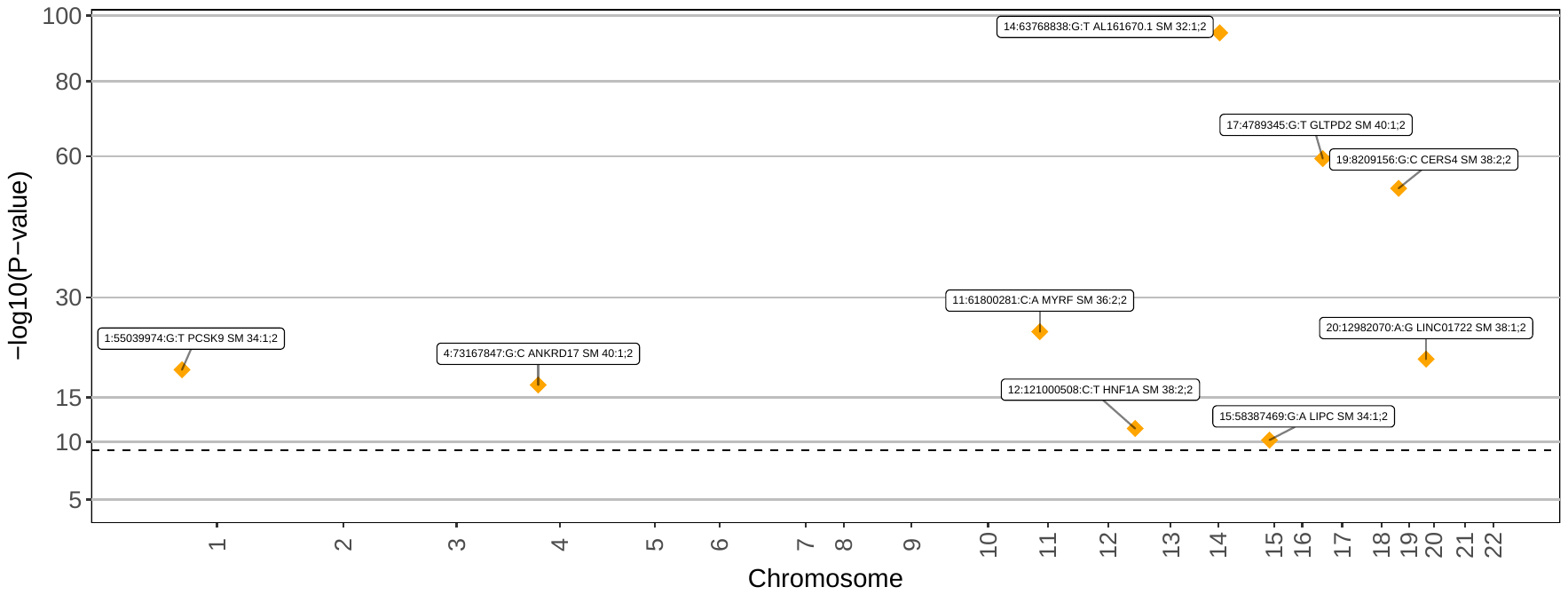

**TAG**

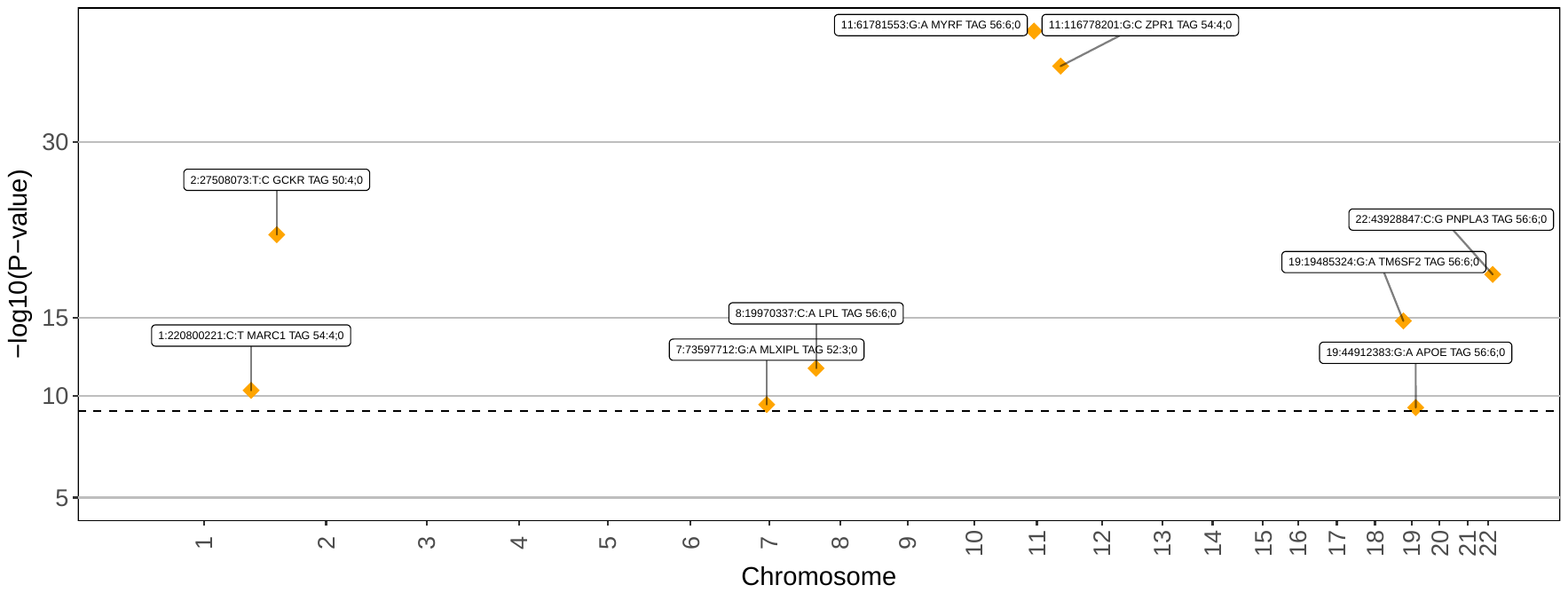

**Manhattan plots for multivariate clusters**

Top associations for signals reaching the Bonferroni-corrected P-value threshold of 4.55e-9 are depicted as diamonds and annotated by variant and locus name. Known loci are colored yellow and novel loci are colored red. The vertical line indicates the Bonferroni-corrected P-value threshold.

**cluster 1**

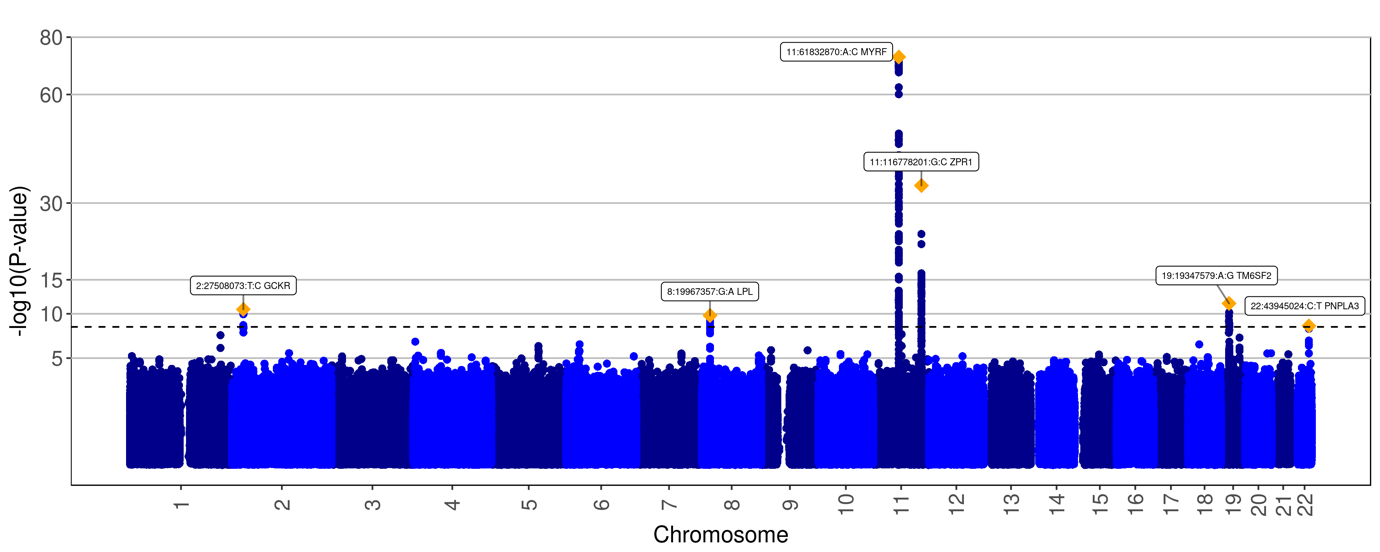

**cluster 2**

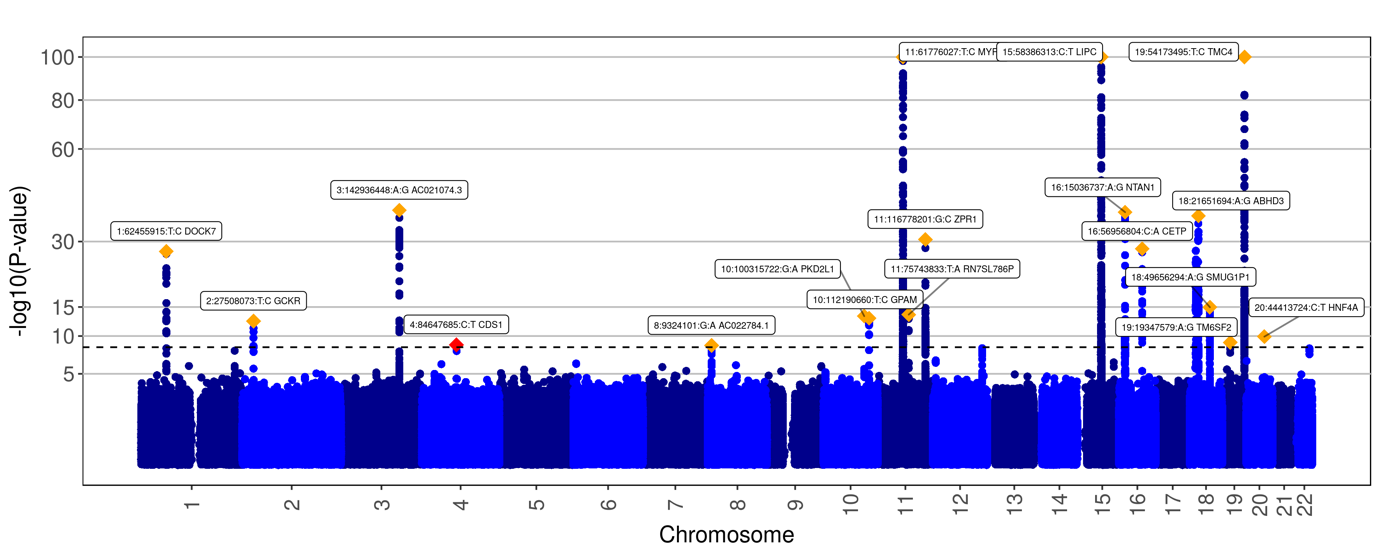

**cluster 3**

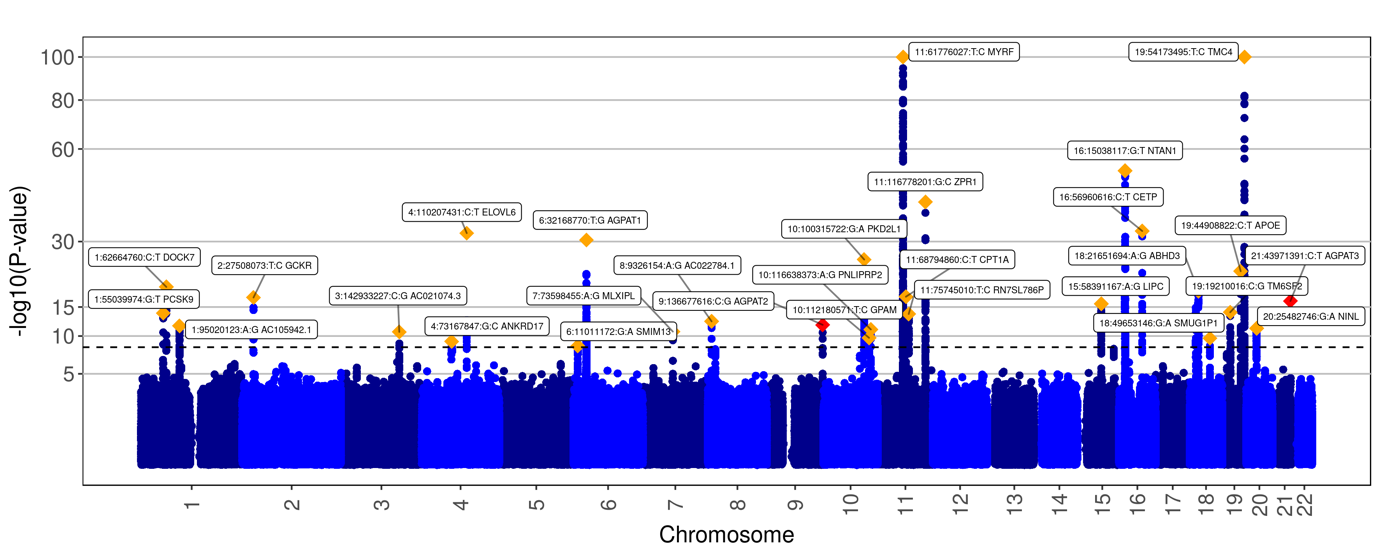

**cluster 4**

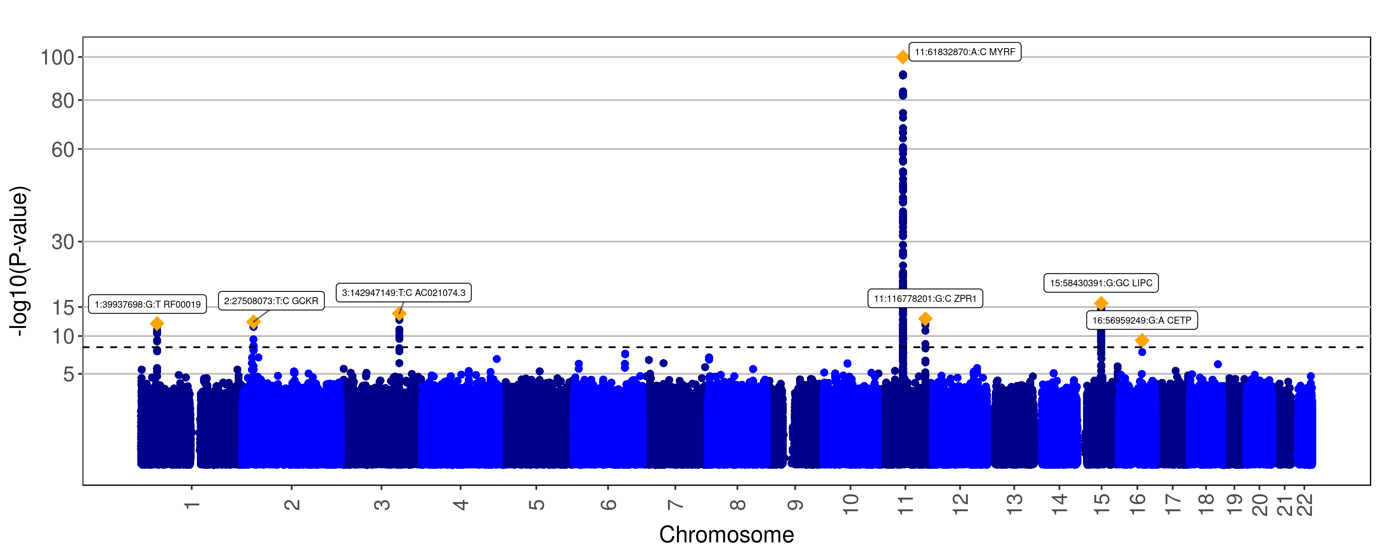

**cluster 5**

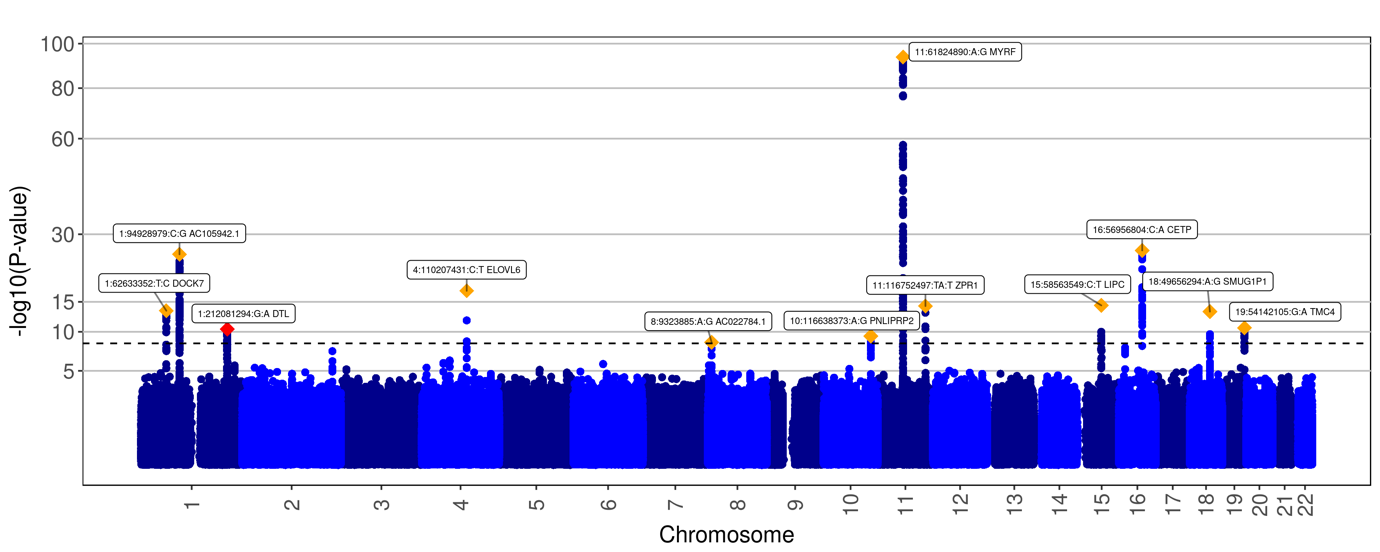

**cluster 6**

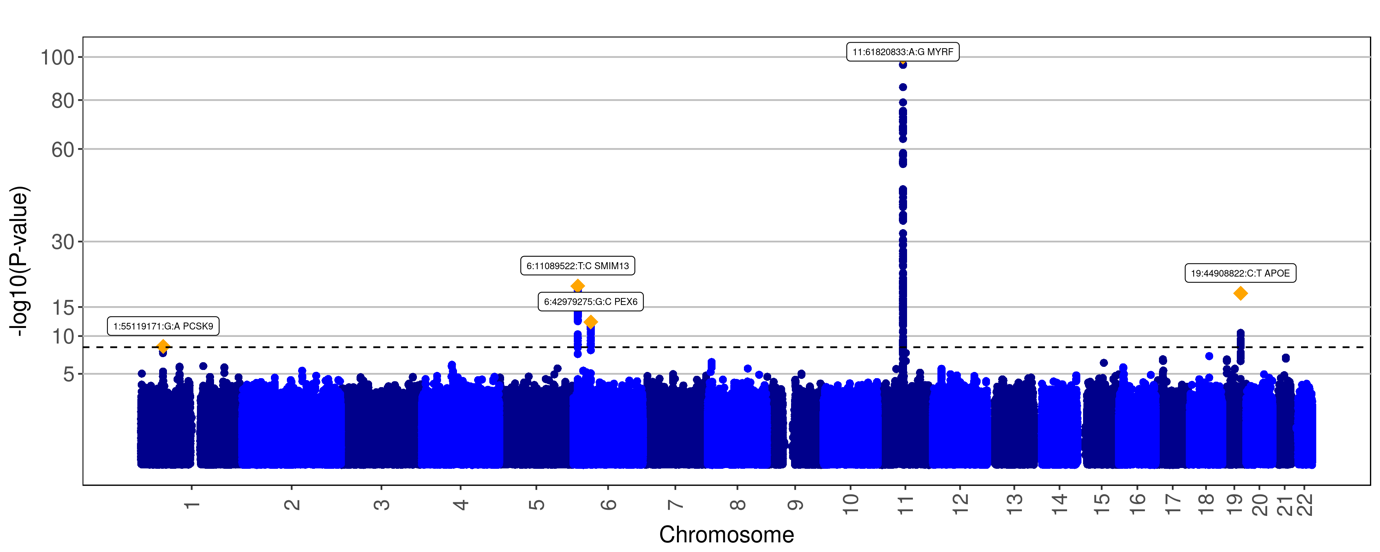

**cluster 7**

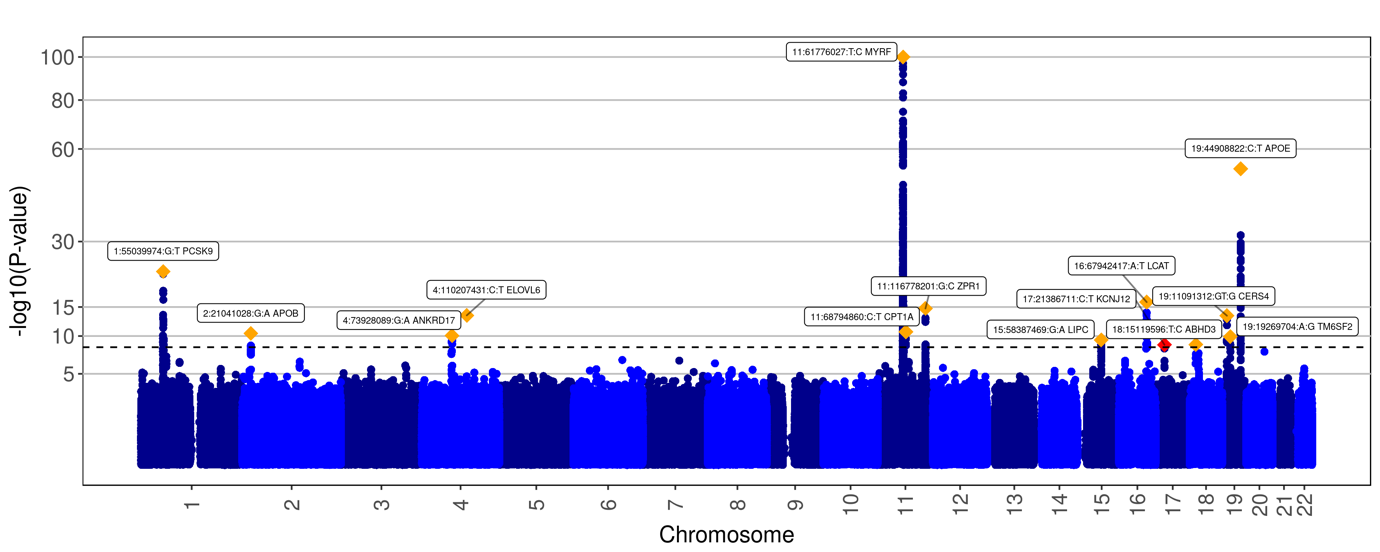

**cluster 8**

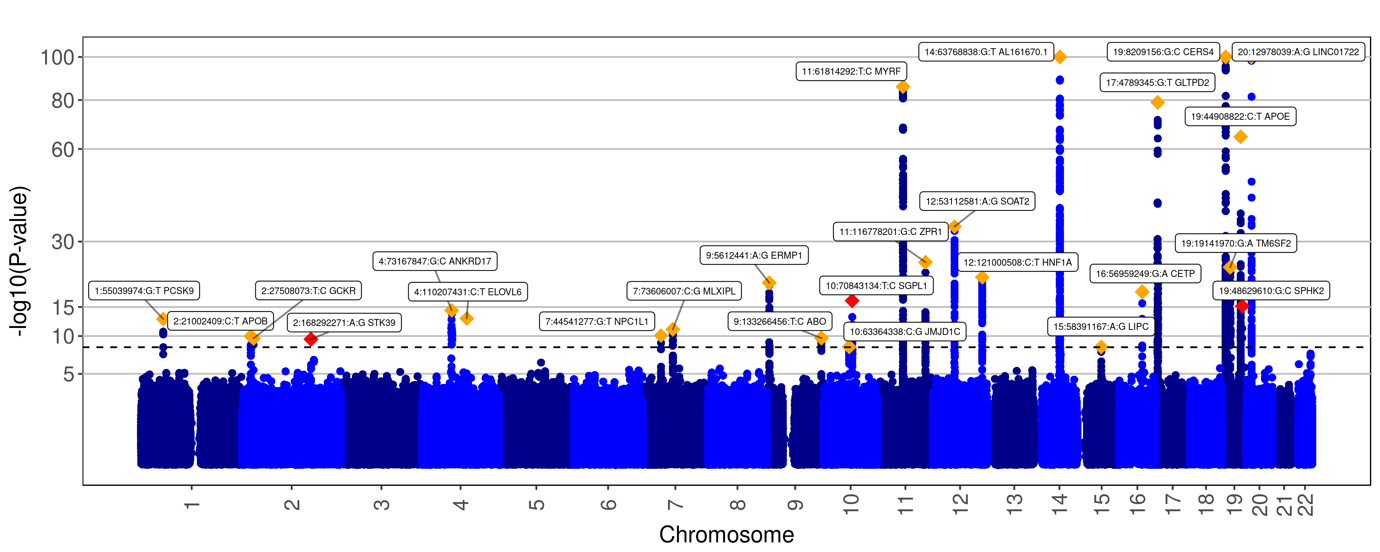

**cluster 9**

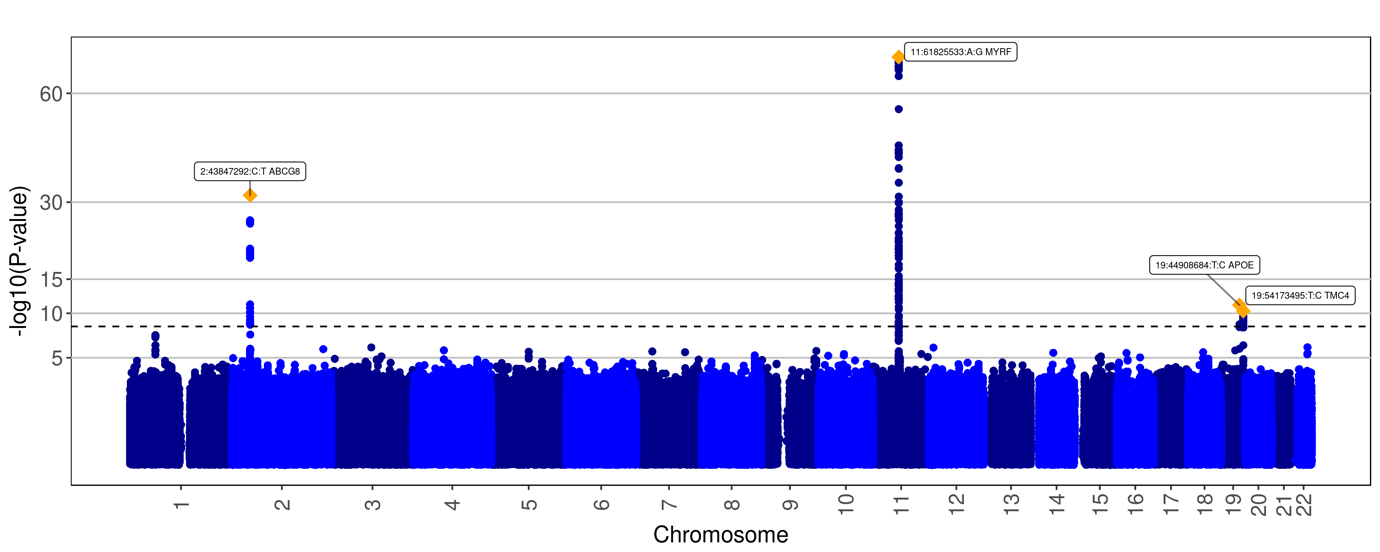

**cluster 10**

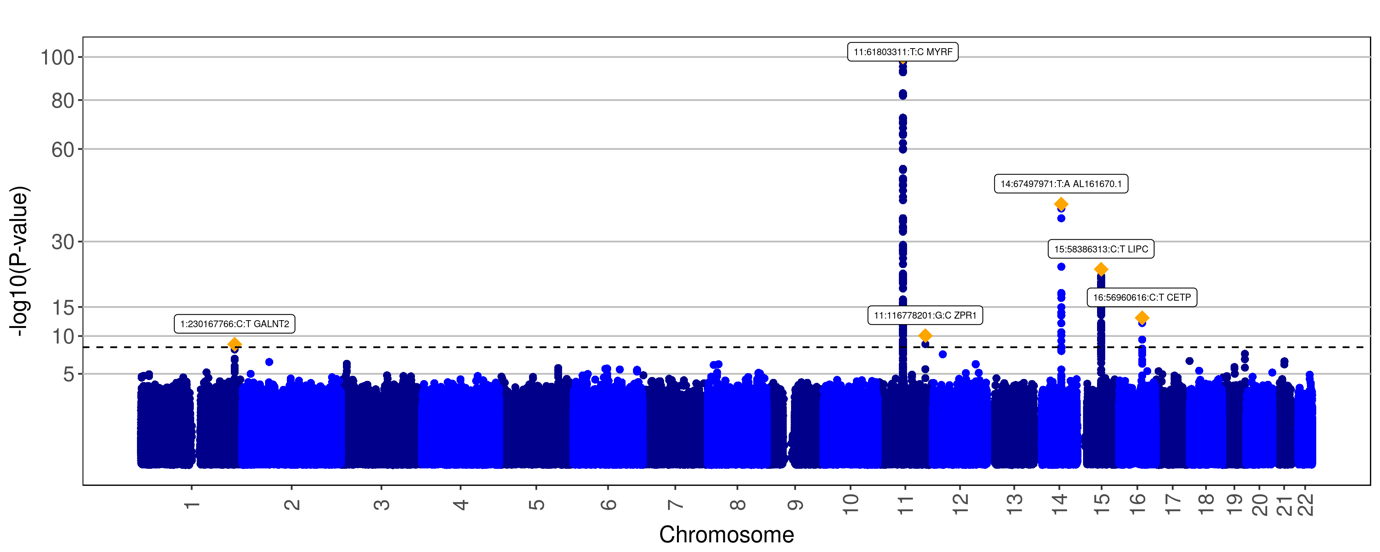

**cluster 11**

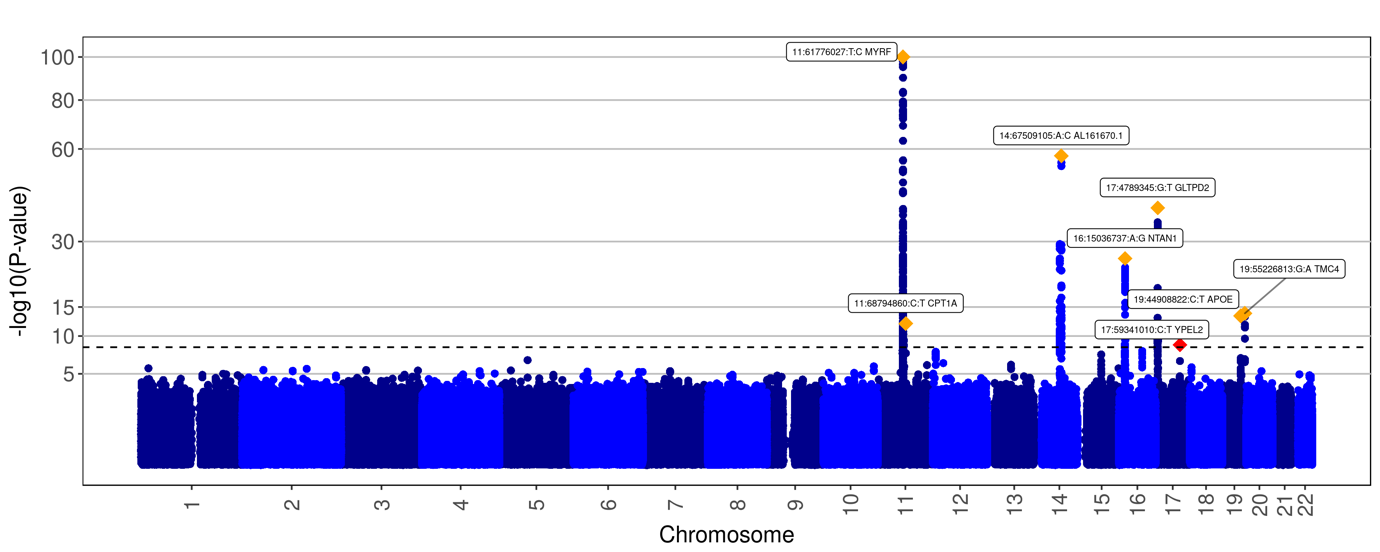
