## Supplementary Note for "Genome-wide association analysis of plasma lipidome identifies 495 genetic associations"

**Simulation of multivariate *P*-values under the null hypothesis**

We simulated multivariate *P*-values for four SNPs with minor allele frequency (MAF) 0.002, 0.012, 0.052 and 0.084 using 100,000 permutations of the genotypes. The simulations were repeated with another set of 100,000 permutations. The QQ-plots showed inflated *P*-values, which are located outside the 95 % confidence interval, for some of the clusters for the rare SNP (MAF < 0.01) and the low-frequency SNP (0.01 $\leq$ MAF < 0.05) and no inflation for the common SNPs (MAF > 0.05).

From the simulations, we conclude that multivariate *P*-values of rare or low-frequency variants can be inflated and therefore we perform these simulations for all rare or low-frequency variants reaching genome-wide significance (GWS) in a multivariate analysis, which did not reach GWS in a univariate GWAS. This was the case for one rare variant and four low-frequency variants.

We then corrected the multivariate *P*-values by the mean of the genomic inflation factors ($\lambda)$, estimated from the simulations as the ratio of the median of the observed chi-square association statistics to its expectation under the null, if the simulations showed inflated *P*-values. More specifically, we divided the$X^{2}$-values by the mean $\lambda$ over the two simulations and recalculated the *P*-values from the corrected $X^{2}$-values. We considered the simulated *P*-values to be inflated if the proportion of *P*-values < 0.01 was greater than 1.2%.

Empirical cumulative distribution function (ECDF)-plots for the five variants are shown in Supplementary Note Figure 1. We summarized the genomic inflation factors, the minimum *P*-value reached in the simulations and the proportion of *P*-values < 0.01 before and after correcting the *P*-values in Supplementary Note Table 1. For the first three variants 6:127321597:T:C, 16:67942417:A:T and 19:48629610:G:C, the proportion of *P*-values < 0.01 was greater than 1.2% and the *P*-values were therefore corrected. The rare variant 6:127321597:T:C is not GWS after the correction and is not reported as a GWS variant.

**Supplementary Note Table 1. Simulation results for rare or low-frequency variants reaching GWS in a multivariate analysis, which did not reach GWS in a univariate GWAS.** Results for both runs of the simulation procedure are shown.

| variant | cluster | MAF | lambda  run1  run2 | P | Corr. P | minP  run1  run2 | Corr. minP  run1  run2 | Prop  P<0.01  run1  run2 | Corr. prop  P < 0.01  run1  run2 |
| --- | --- | --- | --- | --- | --- | --- | --- | --- | --- |
| 6:127321597:T:C | 4 | 0.5% | 1.06  1.05 | 3e-8 | 8e-8 | 2e-6  1e-6 | 3e-6  3e-6 | 1.27%,  1.32% | 1.03%  1.10% |
| 16:67942417:A:T | 7 | 2.8% | 1.06  1.06 | 1e-16 | 8e-16 | 4e-6  4e-6 | 7e-6  8e-6 | 1.21%  1.22% | 0.96%  1.00% |
| 19:48629610:G:C | 8 | 3.1% | 1.06  1.07 | 6e-16 | 5e-15 | 4e-6  6e-7 | 7e-6  1e-6 | 1.21%  1.21% | 0.97%  0.96% |
| 2:168292271:A:G | 8 | 3.1% | 1.05  1.05 | 3e-10 | 8e-10 | 9e-7  4e-6 | 2e-6  7e-6 | 1.14%  1.16% | 0.97%  0.95% |
| 1:212081294:G:A | 5 | 4.2% | 1.04  1.05 | 4e-11 | 1e-10 | 1e-6  2e-5 | 2e-6  3e-5 | 1.09%  1.15% | 0.93%  0.97% |

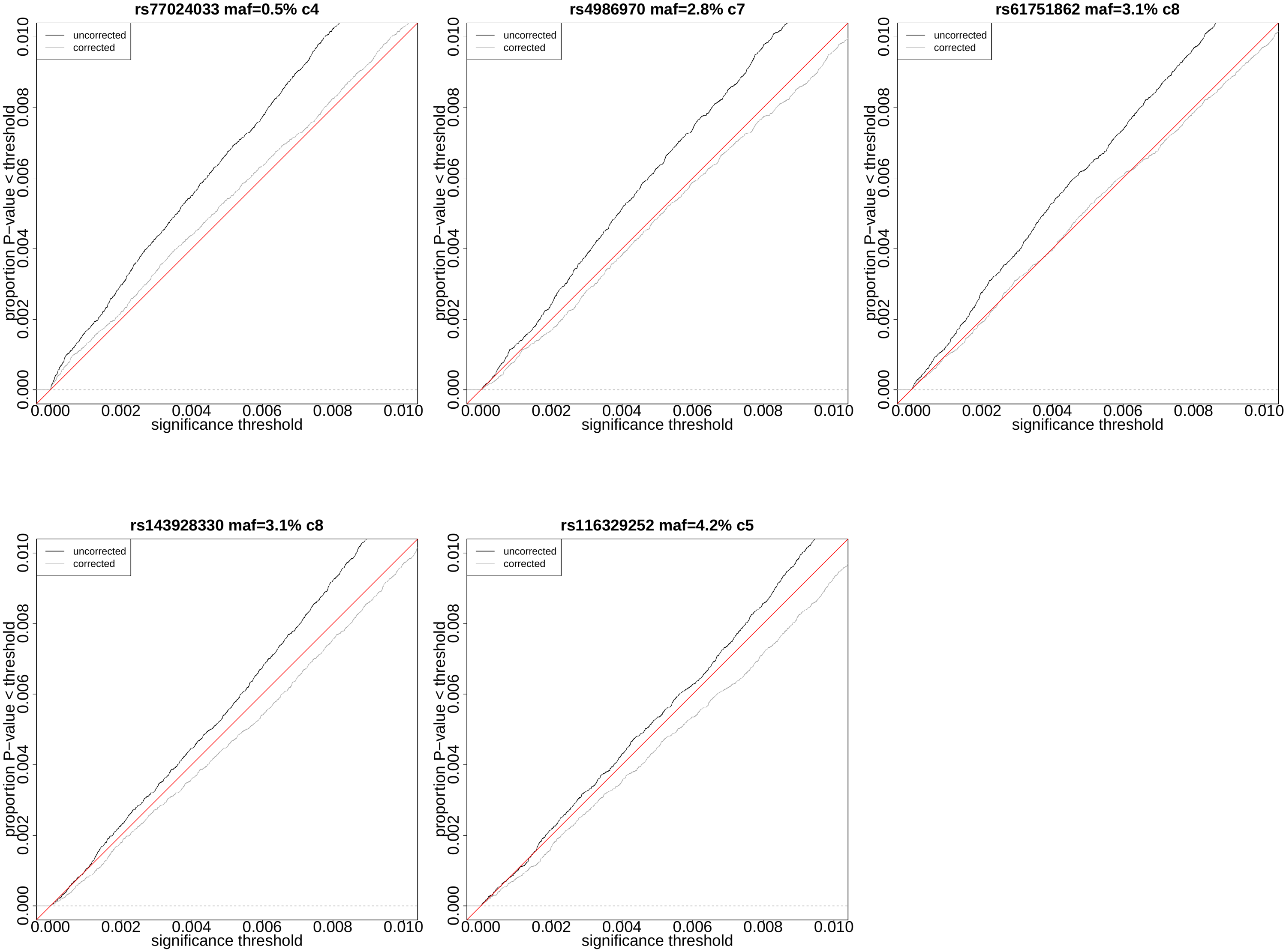

**Supplementary Note Figure 1. Empirical cumulative distribution function (ECDF) plots of simulated *P*-values for rare or low-frequency variants reaching GWS in a multivariate analysis, which did not reach GWS in a univariate GWAS.** The ECDF for uncorrected *P*-values is depicted by a black line and corrected *P*-values by a grey line.

**Rare variants reaching high PIP only in multivariate fine-mapping and low PIP in standard lipids fine-mapping in UKBB**

Three rare variants with CADD score > 10, 15:58541944:G:A (MAF=0.002), 18:21657147:C:T (MAF=0.004) and 6:10995002:C:T (MAF=0.004), reached a PIP > 0.9 in multivariate analysis but not in any univariate analysis and had PIP < 0.001 in standard lipids fine-mapping in UKBB.

The *LIPC* missense variant 15:58541944:G:A reached a PIP of 0.999838 for cluster2 and a *P*-value of 8e-8. The lead SNP of the GWAS region, 15:58386313:C:T, is not rare (MAF=0.338). In univariate fine-mapping the variant is the representative variant for uninformative credible sets of	PE 16:0;0_20:4;0 (PIP=0.9119, *P* =2e-8) and PE 18:0;0_20:4;0 (PIP=0.9217, *P* =3e-8).

The *ABHD3* missense variant 18:21657147:C:T reached high PIPs of 0.9681 and 0.9333 for cluster2 and cluster3. The lead variant of the GWAS region, 18:21651694:A:G, is also rare (MAF=0.007) but reached a low P-value in univariate GWAS of a trait included in cluster2 (*P*=7e-20 for PC 14:0;0_18:2;0) and a trait included in cluster3 (*P*=6e-11 for PC 16:1;0_18:1;0) so no simulation was performed for this variant. The missense variant 18:21657147:C:T reaches *P*-values of 4e-19 and 5e-12 for cluster2 and cluster3, respectively. The variant is a representative variant for PC 14:0;0_18:1;0 (PIP=0.3838, *P*=2e-10) and for an uninformative credible set of PC 14:0;0_18:2;0 (PIP=0.4485, *P*=6e-8). These lipid species are included in cluster2, however the variant was not among the representative variants of traits included in cluster3. The region was only fine-mapped for one trait included in cluster3, PC 16:1;0_18:1;0 (PIP = 0.004, *P*=2e-2).

The *ELOVL2* splice region variant 6:10995002:C:T reached a PIP of 0.9898 and a *P*-value of 2e-7 for cluster6. The lead variant of the GWAS, 6:11089522:T:C, is not rare (MAF=0.153) and reached a *P*-value of 6e-20. The lowest univariate *P*-value of the lead variant for a univariate trait of cluster6 was 1e-5 for CE 22:6;0. The region was not fine-mapped for any univariate GWAS.

**Correlation with standard lipids of traits for which high PIP variants were found by FINEMAP**

We classified the variants for which high PIP variants were found by FINEMAP by their UKB fine-mapping results into 4 categories: (1) variants with CADD score > 10 and PIP$\leq$ 0.1 in UKB, (2) variants with CADD score > 10 and PIP > 0.1 in UKB, (3) variants with CADD score $\leq$ 10 and PIP $\leq$ 0.1 in UKB, (4) variants with CADD score $\leq$ 10 and PIP > 0.1 in UKB.

To compare the correlation between standard lipids and lipid species or LCP-phenotypes, we defined the maximum correlation *mrt* between trait *t* and standard lipids as

$mrt =\max\left( |corr\left( t, HDLC \right)|,|corr\left( t, LDLC \right)|, |corr\left( t,TG \right)|, |corr\left( t,TC \right)| \right)$.

We calculated the maximum correlation *mrv* for a variant *v* as $mrv=\max\left( MRT \right)$, with *MRT* being a list of all *mrt* values for traits *t* associated with *v*. We calculated the mean *mrv* for variants included in each category. The mean *mrv* was lower for variants reaching PIP $\leq$0.1 in UKB (category 1: 0.22, category 3: 0.19) compared to those reaching PIP > 0.1 in UKB (category 2: 0.66, category 4: 0.30). Wilcoxon rank sum test of the two groups had a P-value of 0.01632.

**FinnGen ethics statement**

Patients and control subjects in FinnGen provided informed consent for biobank research, based on the Finnish Biobank Act. Alternatively, separate research cohorts, collected prior the Finnish Biobank Act came into effect (in September 2013) and start of FinnGen (August 2017), were collected based on study-specific consents and later transferred to the Finnish biobanks after approval by Fimea (Finnish Medicines Agency), the National Supervisory Authority for Welfare and Health. Recruitment protocols followed the biobank protocols approved by Fimea. The Coordinating Ethics Committee of the Hospital District of Helsinki and Uusimaa (HUS) statement number for the FinnGen study is Nr HUS/990/2017.

The FinnGen study is approved by Finnish Institute for Health and Welfare (permit numbers: THL/2031/6.02.00/2017, THL/1101/5.05.00/2017, THL/341/6.02.00/2018, THL/2222/6.02.00/2018, THL/283/6.02.00/2019, THL/1721/5.05.00/2019 and THL/1524/5.05.00/2020), Digital and population data service agency (permit numbers: VRK43431/2017-3, VRK/6909/2018-3, VRK/4415/2019-3), the Social Insurance Institution (permit numbers: KELA 58/522/2017, KELA 131/522/2018, KELA 70/522/2019, KELA 98/522/2019, KELA 134/522/2019, KELA 138/522/2019, KELA 2/522/2020, KELA 16/522/2020), Findata permit numbers THL/2364/14.02/2020, THL/4055/14.06.00/2020,,THL/3433/14.06.00/2020, THL/4432/14.06/2020, THL/5189/14.06/2020, THL/5894/14.06.00/2020, THL/6619/14.06.00/2020, THL/209/14.06.00/2021, THL/688/14.06.00/2021, THL/1284/14.06.00/2021, THL/1965/14.06.00/2021, THL/5546/14.02.00/2020, THL/2658/14.06.00/2021, THL/4235/14.06.00/202, Statistics Finland (permit numbers: TK-53-1041-17 and TK/143/07.03.00/2020 (earlier TK-53-90-20) TK/1735/07.03.00/2021, TK/3112/07.03.00/2021) and Finnish Registry for Kidney Diseases permission/extract from the meeting minutes on 4^th^ July 2019.

The Biobank Access Decisions for FinnGen samples and data utilized in FinnGen Data Freeze 9 include: THL Biobank BB2017_55, BB2017_111, BB2018_19, BB_2018_34, BB_2018_67, BB2018_71, BB2019_7, BB2019_8, BB2019_26, BB2020_1, Finnish Red Cross Blood Service Biobank 7.12.2017, Helsinki Biobank HUS/359/2017, HUS/248/2020, Auria Biobank AB17-5154 and amendment #1 (August 17 2020), AB20-5926 and amendment #1 (April 23 2020) and it´s modification (Sep 22 2021), Biobank Borealis of Northern Finland_2017_1013, Biobank of Eastern Finland 1186/2018 and amendment 22 § /2020, Finnish Clinical Biobank Tampere MH0004 and amendments (21.02.2020 & 06.10.2020), Central Finland Biobank 1-2017, and Terveystalo Biobank STB 2018001 and amendment 25^th^ Aug 2020.

**Supplemental acknowledgments**

We want to acknowledge the participants and investigators of FinnGen study. The FinnGen project is funded by two grants from Business Finland (HUS 4685/31/2016 and UH 4386/31/2016) and the following industry partners: AbbVie Inc., AstraZeneca UK Ltd, Biogen MA Inc., Bristol Myers Squibb (and Celgene Corporation & Celgene International II Sàrl), Genentech Inc., Merck Sharp & Dohme LCC, Pfizer Inc., GlaxoSmithKline Intellectual Property Development Ltd., Sanofi US Services Inc., Maze Therapeutics Inc., Janssen Biotech Inc, Novartis AG, and Boehringer Ingelheim International GmbH. Following biobanks are acknowledged for delivering biobank samples to FinnGen: Auria Biobank (www.auria.fi/biopankki), THL Biobank (www.thl.fi/biobank), Helsinki Biobank (www.helsinginbiopankki.fi), Biobank Borealis of Northern Finland (<https://www.ppshp.fi/Tutkimus-ja-opetus/Biopankki/Pages/Biobank-Borealis-briefly-in-English.aspx>), Finnish Clinical Biobank Tampere (www.tays.fi/en-US/Research_and_development/Finnish_Clinical_Biobank_Tampere), Biobank of Eastern Finland (www.ita-suomenbiopankki.fi/en), Central Finland Biobank (www.ksshp.fi/fi-FI/Potilaalle/Biopankki), Finnish Red Cross Blood Service Biobank ([www.veripalvelu.fi/verenluovutus/biopankkitoiminta](http://www.veripalvelu.fi/verenluovutus/biopankkitoiminta)), Terveystalo Biobank ([www.terveystalo.com/fi/Yritystietoa/Terveystalo-Biopankki/Biopankki/](http://www.terveystalo.com/fi/Yritystietoa/Terveystalo-Biopankki/Biopankki/)) and Arctic Biobank (<https://www.oulu.fi/en/university/faculties-and-units/faculty-medicine/northern-finland-birth-cohorts-and-arctic-biobank>). All Finnish Biobanks are members of BBMRI.fi infrastructure ([www.bbmri.fi](http://www.bbmri.fi/)). Finnish Biobank Cooperative -FINBB (<https://finbb.fi/>) is the coordinator of BBMRI-ERIC operations in Finland. The Finnish biobank data can be accessed through the Fingenious^®^ services (<https://site.fingenious.fi/en/>) managed by FINBB.
